# Circulating causal protein networks linked to future risk of myocardial infarction

**DOI:** 10.1101/2025.02.07.25321789

**Authors:** Sean Bankier, Valborg Gudmundsdottir, Thorarinn Jonmundsson, Heida Bjarnadottir, Joseph Loureiro, Lingfei Wang, Nancy Finkel, Anthony P Orth, Thor Aspelund, Lenore J Launer, Johan LM Björkegren, Lori L Jennings, John R Lamb, Vilmundur Gudnason, Tom Michoel, Valur Emilsson

## Abstract

Variations in blood protein levels have been associated with a broad spectrum of complex diseases, including atherosclerotic cardiovascular disease (ACVD). These associations highlight the intricate interplay between local (e.g., cardiovascular) and systemic (non-cardiovascular) factors for the development of ACVD, emphasizing the need for a comprehensive, systems-level understanding of its etiology. To accomplish this, we developed a causal network inference framework by analyzing one of the largest serum proteomics studies to date, the Age, Gene/Environment Susceptibility-Reykjavik Study (AGES), a prospective population-based study of 7,523 serum proteins measured in 5,376 older adults. To reconstruct a causal network of serum proteins, we used *cis*-acting protein quantitative trait loci (pQTLs) as instrumental variables to infer causal relationships between protein pairs, while accounting for potential unobserved confounding factors. We identified 185 causal protein subnetworks (FDR = 1%, n ≥ 10 members), which collectively interacted with 5,611 target proteins, offering valuable biological insights and an overview of systemic homeostasis. Several subnetworks, many of which interact to establish a hierarchy of directional relationships, were significantly associated with future myocardial infarction and/or its long-term complications like heart failure, as well as with key cardiometabolic traits that contribute to the onset of ACVD.

## Introduction

Atherosclerotic cardiovascular disease (ACVD) is the leading cause of age-standardized deaths globally^1^. While lipid-lowering treatments have been shown to reduce the risk of ACVD^2^, residual risk persists^3–5^, underscoring a significant and unmet medical need. Early coronary atherosclerosis, which advances to coronary artery disease (CAD), is the primary cause of ACVD. In its most advanced stage, coronary artery plaques may rupture, manifesting clinically either through myocardial infarction (MI) or stroke. The rate of coronary plaque growth is influenced by various systemic factors across multiple organ systems, such as the immune system driving systemic inflammation, the liver regulating lipoprotein metabolism, and adipose tissue and skeletal muscle contributing to the development of obesity and type 2 diabetes (T2D)^6^. Other contributing factors are endocrine signaling and hemodynamic processes^6^. The rate at which CAD progresses depends on the interplay between these systemic factors, ultimately leading to plaque rupture, thrombus formation, and MI^7,6^. The complex etiology of ACVD is shaped by both genetic and lifestyle factors, which are mediated through interactions between multiple organ systems. The molecular disruptions across organs that contribute to ACVD have primarily been studied in isolated pathways using model systems. While these studies offer valuable insights into disease etiology and treatment, they fail to capture the systemic complexity of ACVD. In other words, the rate at which ACVD progresses to become clinically significant depends on the interplay of both local (cardiovascular) and systemic (non-cardiovascular) factors, an aspect of the disease that is often overlooked. Thus, a broader systems-level approach is required to obtain a more wholistic understanding of its etiology.

Tissue-specific and cross-tissue transcriptional networks, have been established as both undirected networks^8–12^ and directed networks^13,14^, operating within and across various tissues, and demonstrating robust associations with complex diseases. In contrast to undirected co-regulatory networks, directed networks have the potential to differentiate between causal and reactive nodes and elucidate how these causal nodes propagate their effects^15^. Gene expression quantitative trait loci (eQTLs) have been employed as genetic instruments in causal inference analysis and as priors in reconstructing transcriptional Bayesian networks^13^. These variants offer an effective means of natural perturbation, to infer causal relationships between genes and higher-order phenotypes like disease, and even between genes themselves^10,12^. This has been well documented by the explosion of Mendelian Randomization (MR) studies that use genetic variants as instrumental variables for both molecular and higher order phenotypes^16^. Although reconstructing directed causal networks has traditionally been computationally intensive and limited to small-scale systems, recent advancements have significantly enhanced performance^14,17–19^. These improvements have made the process orders of magnitude faster, enabled the coverage of a larger proportion of variance in the data, and proven especially effective when both genotype and molecular node data are available for the same sample^17,18^.

Proteomics has recently advanced to the point where high-throughput measurements allow for the analysis of thousands of proteins from a single tissue or biofluid sample in large population studies^20,21^, exposing the depth and complexity of the plasma and serum proteomes. These recent advancements have largely been driven by the highly sensitive aptamer-based affinity methods. In fact, comparisons between various proteomics platforms highlight the superior performance of the aptamer-based platform, especially in terms of detection precision and sensitivity^22^. Using this approach led to the identification of the first human protein co-regulatory network, reconstructed from the analysis of 4,137 serum proteins measured in 5,457 older adults of the prospective, population-based AGES cohort^21^. Furthermore, we demonstrated that the network modules for circulating proteins are under strong genetic control and are linked to a broad range of past, current and future disease states^21^. Notably, the structure and composition of these serum protein networks are stable and have been validated in other body fluids, such as cerebrospinal fluids^23^. Finally, unlike solid tissue networks, the serum protein network consists of modules incorporating proteins synthesized by many, if not all, tissues across the body^21,24^.

In this study, we describe the reconstruction of a directed circulating Causal Protein Network (CPN) within the AGES cohort using an expanded dataset comprising 7,523 serum proteins encoded by 6,586 genes, with *cis*-acting protein quantitative trait loci (pQTLs) serving as instrumental variables. Applying various filters, including network size and genome linkage disequilibrium between instrumental variables, we identified 185 CPN subnetworks, encompassing at least 10 protein members and interacting with a total of 5,611 target serum proteins. The CPN subnetworks were examined for their relationships with each other and tested for associations with various ACVD related outcomes. We highlight the subnetworks with the strongest associations to new-onset MI, along with cardiometabolic traits that contribute to the risk of ACVD.

## Results

### Study population and analysis overview

This study builds on the population-based prospective AGES cohort of older adults (N = 5,764, mean age 76.6 ± 5.6 years, age range 66-96 years, 57% female). The cohort is richly annotated with data on disease risk factors, endpoints, comorbidities, genotype information, and deep serum proteomics^21,25^. Table 1 displays the demographic, biochemical, clinical, physiological, and anthropometric data, as well as cardiovascular imaging measurements of participants in the AGES study, analyzed for 7,523 serum proteins, and stratified by incident MI (n = 668) after excluding all prevalent MI cases. The follow-up period for newly diagnosed MI patients extended up to 12 years from baseline, with recurrent cases excluded from the incident group analysis. Person-years of follow-up were calculated from the first AGES visit until the date of diagnosis, death, or the end of the follow-up period, with a median of 5.35 [2.59, 8.16] years for the incident MI group (Table 1). As anticipated, several measures associated with an increased risk of an MI event are significantly altered in the incident MI group compared to the non-MI group (Table 1). These include a higher prevalence of metabolic syndrome (MetS) and type 2 diabetes (T2D), as well as elevated coronary artery calcium (CAC) and carotid artery plaque burden (Table 1). Furthermore, 25.7% of the incident MI group developed heart failure (HF), compared to 4.6% of the non-MI group (P < 0.001) (Table 1). Figure 1 presents an overview of the study and its workflow, including the reconstruction of the circulating CPN within the AGES study, as well as key disease endpoints and cardiometabolic traits associated with ACVD that are examined in the present study. Additional details on the data and analyses are provided in Supplementary Figure S1.

**Figure 1.**
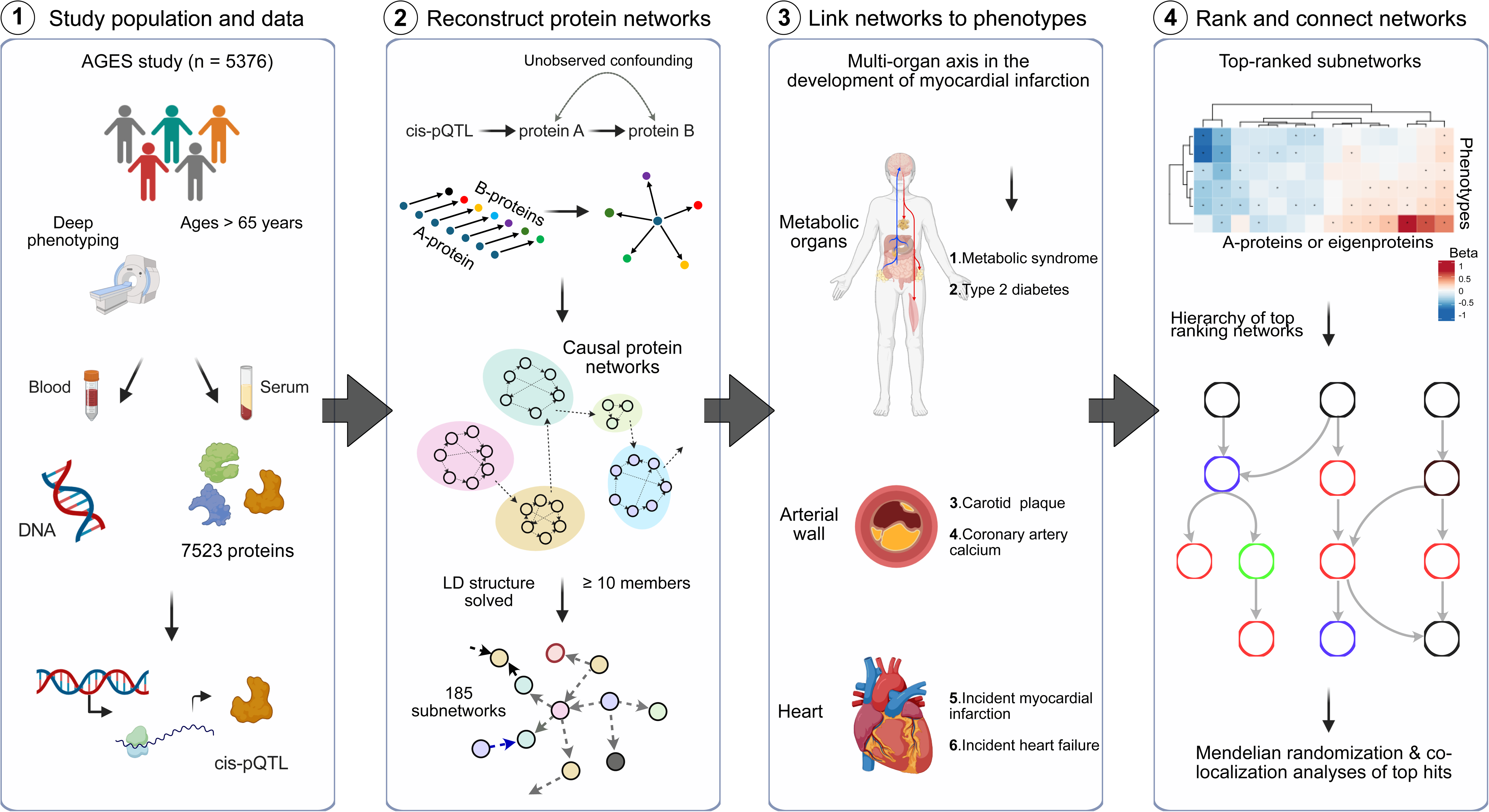
A flowchart outlining the study overview and approach. The figure illustrates the study steps (1-4), starting with 1) the well-annotated, prospective AGES population-based study, which includes comprehensive phenotype and genotype data, extensive serum proteomic measurements, and the identification of *cis*-acting protein quantitative trait loci (pQTLs). 2) The reconstruction and identification of 185 circulating causal protein networks (CPNs), each comprising 10 or more targets, were achieved using a 1% false discovery rat (FDR) threshold based on the principles of Mendelian Randomization (MR) analysis. 3) The association analysis of both network regulators and network eigenproteins with phenotypes that reflect activity in key organ systems involved in atherosclerotic cardiovascular disease (ACVD). 4) The rank-ordering of CPN subnetworks based on the number of links to new-onset myocardial infarction (MI) and related phenotypes, showing how these subnetworks form a hierarchical structure.

**Table 1.** Baseline characteristics of the AGES study participants, stratified by incident myocardial infarction (MI). All participants were measured for 7,523 proteins in serum. The table was generated after removing all prevalent MI cases.

| Characteristic | Variable* | Free of MI | Incident MI | P-value | Total |
| --- | --- | --- | --- | --- | --- |
| **Demographics** |  |  |  |  |  |
|  | Numbers | 4322 (86.6) | 668 (13.4) | N/A | 4990 |
|  | Females (%) | 2690 (62.2) | 335 (50.1) | < 0.001 | 3025 (60.6) |
|  | Age (years) | 76.53 (5.78) | 79.22 (6.04) | < 0.001 | 76.89 (5.88) |
| **Anthropometry** |  |  |  |  |  |
|  | BMI (kg/m<sup>2</sup>) | 27.05 (4.50) | 26.98 (4.25) | 0.724 | 27.04 (4.47) |
|  | BMI category |  |  | 0.517 |  |
|  | BMI < 25 kg/m<sup>2</sup> | 1450 (33.9) | 214 (32.5) |  | 1664 (33.7) |
|  | BMI = 20-30 kg/m<sup>2</sup> | 1861 (43.5) | 302 (45.9) |  | 2163 (43.8) |
|  | BMI ≥ 30 kg/m<sup>2</sup> | 966 (22.6) | 142 (21.6) |  | 1108 (22.5) |
|  | SAT (cm<sup>2</sup>, CT) | 260.85 (113.31) | 245.53 (105.12) | 0.002 | 258.85 (112.38) |
|  | VAT (cm<sup>2</sup>, CT) | 169.52 (78.65) | 180.21 (83.53) | 0.002 | 170.91 (79.38) |
| **Lifestyle** |  |  |  |  |  |
|  | Smoker |  |  | 0.066 |  |
|  | Never | 1946 (46.5) | 267 (41.7) |  | 2213 (45.8) |
|  | Former | 1747 (41.7) | 285 (44.5) |  | 2032 (42.1) |
|  | Current | 496 (11.8) | 88 (13.8) |  | 584 (12.1) |
| **Physiological** |  |  |  |  |  |
|  | DBP (mmHg) | 79.83 (10.75) | 80.65 (10.82) | 0.068 | 79.94 (10.76) |
|  | SBP (mmHg) | 151.02 (22.41) | 156.07 (24.18) | < 0.001 | 151.70 (22.71) |
|  | eGFR (ml/min/1.73m<sup>2</sup>) | 64.80 (15.23) | 59.32 (16.28) | < 0.001 | 64.07 (15.48) |
| **Cardiovascular imaging** |  |  |  |  |  |
|  | CAC | 209.3 [26.9, 689.8] | 670.4 [192.8, 1611.1] | < 0.001 | 241.1 [35.8, 792.3] |
|  | TAC | 204.8 [21.0, 828.7] | 556.1 [97.3, 1550.5] | < 0.001 | 234.0 [26.9, 914.4] |
|  | Plaque severity | 2473 (64.2) | 455 (79.0) | < 0.001 | 2928 (66.1) |
| **Metabolic** |  |  |  |  |  |
|  | T2D | 499 (11.5) | 105 (15.7) | 0.003 | 604 (12.1) |
|  | MetS | 847 (19.6) | 158 (23.7) | 0.017 | 1005 (20.1) |
|  | HbA1c | 0.48 (0.09) | 0.50 (0.10) | < 0.001 | 0.49 (0.09) |
|  | Incident NAFLD | 38 (5.6) | 10 (10.6) | 0.093 | 48 (6.2) |
|  | HDLc (mmol/L) | 1.62 (0.45) | 1.52 (0.44) | < 0.001 | 1.60 (0.45) |
|  | LDLC (mmol/L) | 3.75 (0.95) | 3.80 (1.00) | 0.155 | 3.75 (0.96) |
|  | TG (mmol/L) | 0.05 [-0.24, 0.36] | 0.10 [-0.23, 0.46] | 0.005 | 0.05 [-0.24, 0.38] |
| **Medication** |  |  |  |  |  |
|  | Lipid lowering | 713 (16.5) | 151 (22.6) | < 0.001 | 864 (17.3) |
|  | Antihypertensive | 2557 (59.6) | 478 (72.4) | < 0.001 | 3035 (61.3) |
| **Cardiovascular and follow-up** |  |  |  |  |  |
|  | Incident CHD | 402 (9.3) | 518 (77.5) | < 0.001 | 920 (18.4) |
|  | Incident HF | 198 (4.6) | 172 (25.7) | < 0.001 | 370 (7.4) |
|  | Incident Stroke | 395 (9.1) | 56 (8.4) | 0.574 | 451 (9.0) |
|  | Follow-up (years) | 9.86 [7.47, 10.94] | 5.35 [2.59, 8.16] | < 0.001 | 9.66 [6.24, 10.79] |

\*Numbers are mean (SD) for continuous-, N (%) for categorical- and median [IQR] for skewed variables. The LDL cholesterol and blood pressure readings were adjusted for lipid-lowering and antihypertensive medications, respectively. The reported P-values are two-sided. Abbreviations: BMI, body mass index; SAT, subcutaneous fat by CT; VAT, visceral fat by CT; SBP, systolic blood pressure; DBP, diastolic blood pressure; HDLC, HDL cholesterol; LDLC, LDL cholesterol; TG, triglyceride; FG, fasting blood glucose; HbA1c, glycated hemoglobin T2D, type 2 diabetes; MetS, metabolic syndrome; NAFLD, non-alcoholic fatty liver disease; CHD, coronary heart disease; MI, myocardial infarction; HF, heart failure; CAC, coronary artery calcium; TAC, thoracic aortic calcium; Plaque, carotid plaque severity (carotid plaque was assessed in 5017 individuals of the AGES cohort); N/A, not applicable.

### Reconstruction of the circulating causal protein network

We reconstructed a global network of circulating proteins using a causal inference framework, inferring edges from pairwise protein relationships, with *cis*-acting pQTLs at a false discovery rate (FDR) < 5% serving as instrumental variables (Methods). A total of 5,662 proteins (aptamers) with a *cis*-acting pQTL instrument, referred to as A-proteins (Figure 1), were identified. For each A-protein, we calculated the probability of it having a causal effect on the serum levels of each of the remaining 7,523 proteins, referred to as B-proteins or targets, yielding approximately 42.5 million potential network edges. From all highly significant edges (FDR = 1%), we defined each network regulator and its targets as a subnetwork of the global CPN. At this stage, nearly half of all network regulators had only a single target and at more permissive FDR thresholds, the proportion of A-protein with just one target decreases further (Supplementary Figure S2). Since our primary focus was on regulatory proteins that accounted for the most variation in the serum proteome, we selected A-proteins with a minimum of 10 targets, leading to a global network consisting of 43,528 edges and 234 A-proteins. For A-proteins with multiple aptamers, we selected those with the largest number of targets, and further refinement of linkage disequilibrium (LD) among *cis*-acting instruments resulted in the final CPN comprising 185 A-proteins (referred to here as network regulators), their corresponding subnetworks, and a total of 31,358 edges (Supplementary Table S1, Figure 2A). We identified a high number of indirect regulations (Figure 2A), where two network regulators (*x* and *y*) are responsible for the regulation of a common set of targets (*z*), but *y* is also a target of *x*. Such motifs are known as feed-forward loops (FFLs) and are a common feature in biological networks^26^.

**Figure 2.**
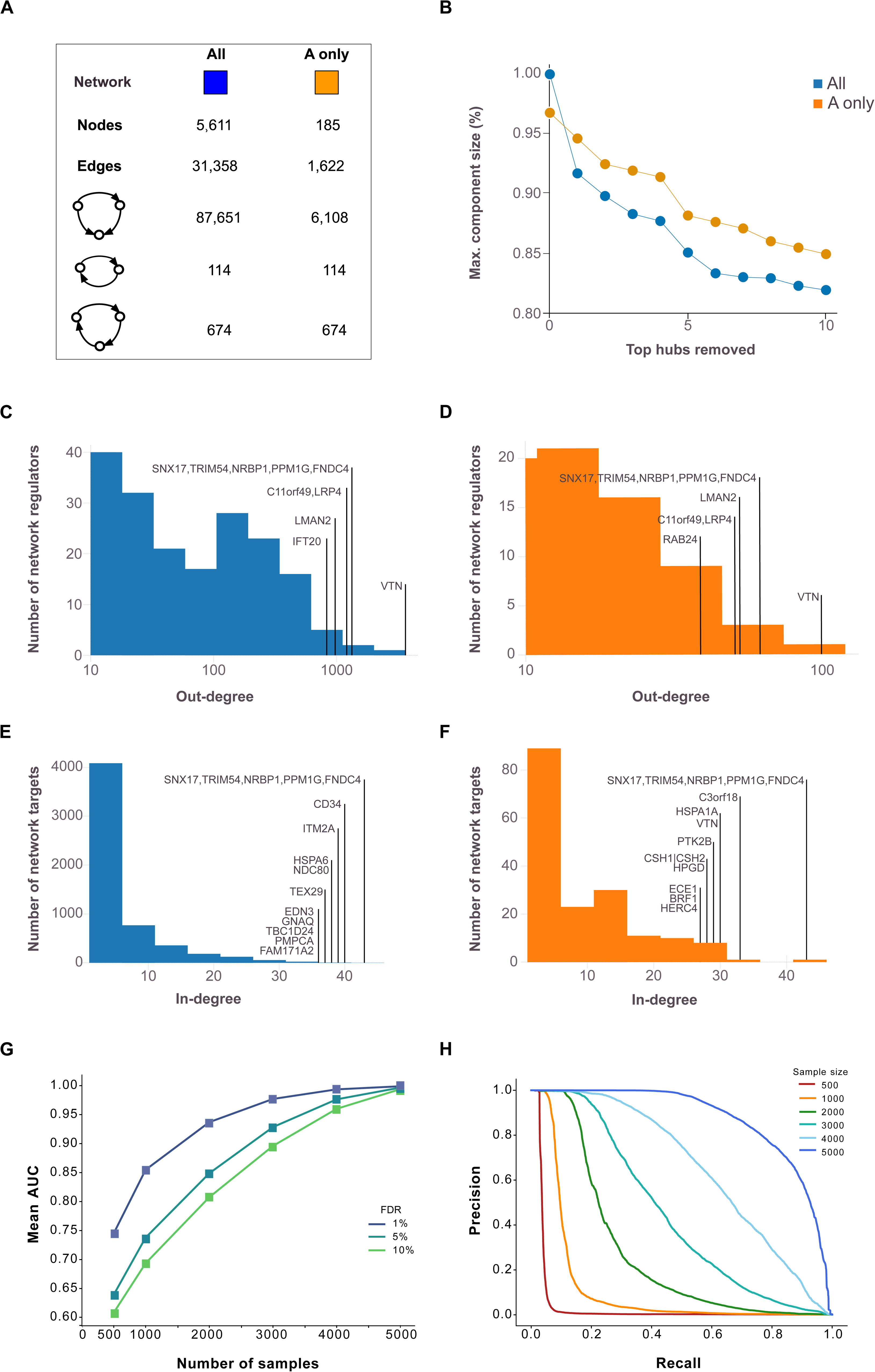
Analysis of degree distribution and robustness in causal protein networks. The figure shows the basic statistics for both the complete causal protein network (CPN) (all) and the restriction to edges between the 185 causal protein network regulators containing at least 10 members (A-only), including the number of nodes, edges, and 2/3-node feedback and feedforward loops. (**B**) shows the relative sizes of the largest connected component after deleting the k=1 to 10 top ‘hub’ proteins. (**C**) Out-degree distributions for network regulators are presented for the complete network (blue) and (**D**) the A-only network (orange), with annotations emphasizing the most highly connected network regulators. (**E**) The corresponding in-degree distribution is shown for the network target B-proteins for the complete network (blue) and (**F**) A-only network (orange). (**G**) Robustness analysis receiver operating characteristic (ROC) area under the curve (AUC) for sub-sampled networks compared to networks using all 5,376 samples. (**H**) Precision-Recall curves for sub-sampled networks at a 1% FDR threshold are compared to networks using all samples. Similar curves at 5% and 10% FDR thresholds are presented in Supplementary Figure S4.

Independent *cis* instruments accounted for an average of 7.4% of the variance in protein expression across networks, with some *cis* signals contributing up to 84% of the variance (Methods). Furthermore, by utilizing *cis* signals for parental nodes (Methods, Supplementary Text), we observed a correlation between the number of regulators a target protein has, and the proportion of total variance in target protein expression explained by the *cis* signal of the network regulator (Spearman r = 0.78) (Supplementary Figure S3A-B). In some cases, more than 50% of the variance in the expression of the target protein was explained solely by the *cis*-acting pQTLs of the parent proteins, with no contribution from local *cis*-components (Supplementary Text).

### High robustness and edge precision in the circulating causal protein network

We found the CPN to be robust in response to hub removal (Figure 2B), with the largest connected component size still containing more than 80% of all nodes following the removal of the top 10 largest hubs, suggesting high levels of co-regulation and biological redundancy. The CPN also exhibited the typical “scale-free” property of biological networks^27^, where a small number of regulators have a very high number of targets (Figure 2C-D). We also examined the distribution of incoming edges and found that many of the network regulators with a high number of outgoing edges also have many incoming edges, further highlighting the interconnectedness of the global CPN (Figure 2E-F).

Network robustness analysis and Precision-Recall assessments for the networks were conducted across various FDR thresholds and sample sizes. This involved random sampling of AGES individuals at varying sample sizes (Methods) and demonstrated a mean area under the receiver operating characteristic (ROC) curve exceeding 90%, even when the sample size was reduced to 2,000 (Figure 2G). However, when represented as a Precision-Recall curve, the precision at an FDR of 1% declined sharply to higher recall levels for sample sizes below 3,000 and was even lower at more permissive FDR thresholds (Figure 2H, Supplementary Figure S4). Lastly, pairwise correlations between network targets were significantly stronger (Kruskal-Wallis P-value < 10^-300^) compared to randomly selected protein groups of the same size (Supplementary Figure S5). In summary, these findings emphasize the strengths of the AGES study’s large sample size, highlighting the robustness and accurate edge estimation of the identified protein network structure.

### Hierarchical organization of the circulating causal protein network

To introduce acyclicity to the 1,622 interactions among all network regulators, we used a greedy heuristic to analyze the structure of the CPN and establish the hierarchy among the network regulators (Methods). This process led to the removal of 572 edges between network regulators, resulting in an acyclic network with 1,050 interactions, and the nodes then arranged at different levels according to their distance from the roots of the network (Figure 3). We then reintroduced the removed edges to the CPN, while maintaining the ordered layout, and found much of the hierarchical structure to be conserved within the different levels (Supplementary Figure S6). Most edges between proteins were observed within single levels, either above or below, with a few crossing multiple levels, indicating a highly structured organization within these networks. A hierarchical structure with a small number of levels has been observed in other biological networks^27,28^.

**Figure 3.**
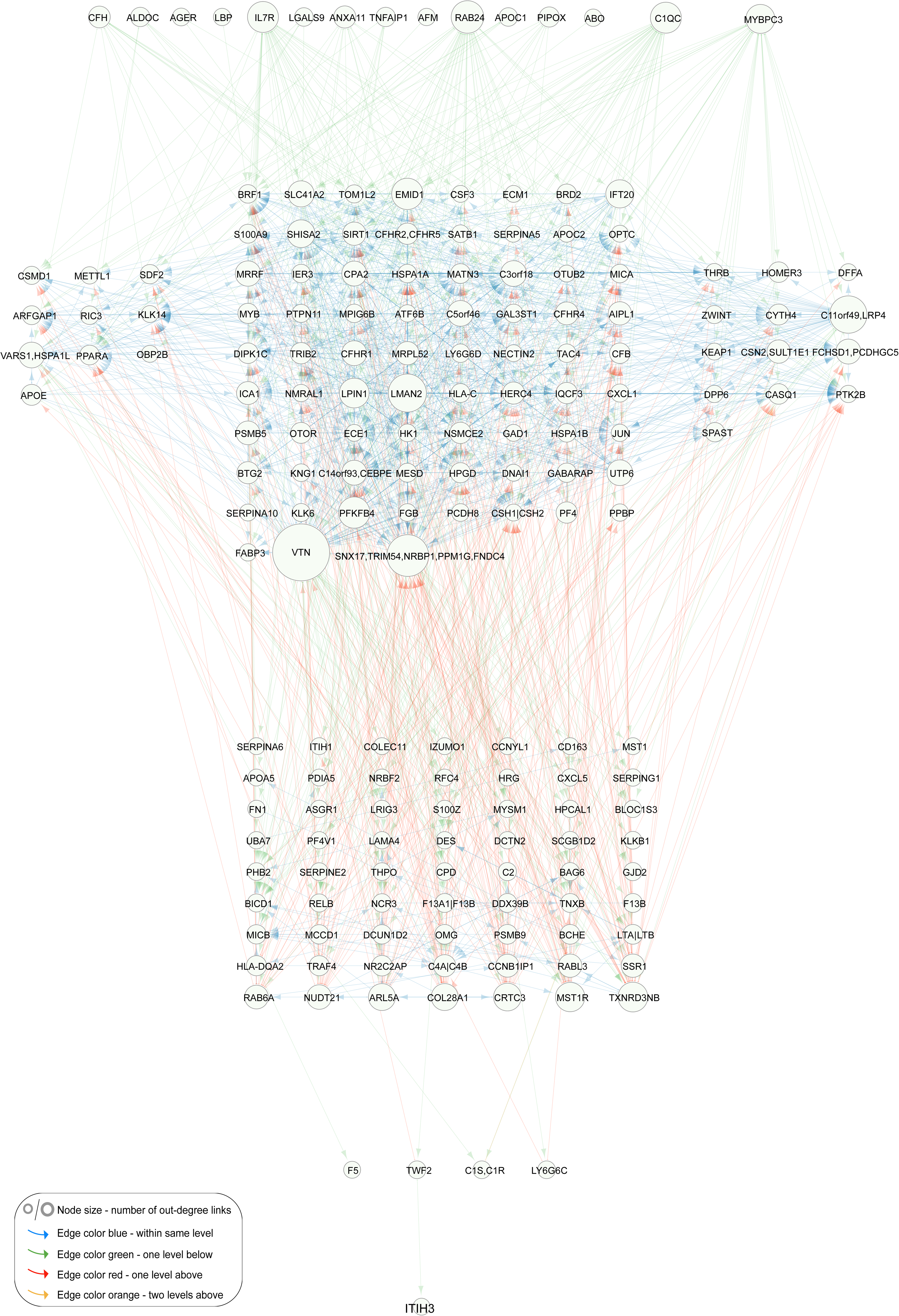
Interactions between network regulators of the circulating causal protein network. Network regulator only directed acyclic graph (DAG) of the circulating causal protein network (CPN). Network visualization of causal interactions between the network regulators of the 185 subnetworks with 10 or more targets (FDR = 1%). Node size is proportional to the number of out-degree links. The edges are color-coded according to levels (hops): blue for connections within the same level, green for links to one level below, red for links to one level above, and orange for links to two levels above. A similar network visualization is presented in Supplementary Figure S6, where no edges have been removed.

Our approach infers causal relationships between protein pairs (A→B), whether direct or indirectly mediated by a third variable (e.g., A→C→B). The abundance of FFLs and within-level interactions may therefore reflect the high sensitivity of the AGES data for detecting weaker indirect causal effects. To address this, we generated a version of the directed acyclic graph (DAG) with all indirect edges removed, known as the transitive reduction of the DAG (Methods). This simplified, cascade-like network, comprising 255 edges, showed that many previously highly interconnected nodes remained prominent even after indirect edges were excluded (Supplementary Table S2), further highlighting the ability of the AGES study to uncover indirect causal effects within the hierarchical organization of the CPN.

Protein-protein interaction (PPI) networks are highly modular, and feature connected hub proteins, reflecting a hierarchical biological organization^29^. Using the human integrated protein-protein interaction reference database^30^, we identified edges in the serum protein network, which overlap with direct physical PPI networks, as outlined in the Supplementary Text (Supplementary Material). Specifically, across all confidence thresholds, we found that the true CPN captured significantly more PPI edges than the average observed in random networks, including the highest confidence PPIs (z-score = 24.3, P-value <0.001) (Method, Supplementary Text, Supplementary Figure S7). This comparison highlights that a substantial portion of the CPN may reflect direct physical interactions, most of which have been identified *in vitro* from solid tissues^30^. However, unlike the CPN, the PPI network database cannot capture interactions that span tissue boundaries within the context of the serum.

### Causal protein networks linked to ACVD related outcomes

We assessed the association of each network regulator and its corresponding CPN eigenprotein with incident MI and ACVD-related cardiometabolic traits, including MetS, T2D, CAC, carotid artery plaque burden, and incident HF (Methods). For this analysis, the first principal component (PC1) of each of the 185 subnetworks was calculated, and PC1s accounting for more than 15% of the variance within their respective subnetworks were designated as eigenproteins. Three CPN subnetworks did not meet this criterion and, therefore, did not have valid eigenproteins. Overall, 50 network regulators and 36 eigenproteins were significantly associated with incident MI (Supplementary Tables S3 and S4). Figure 4 illustrates the correlation between network regulators and their corresponding eigenproteins, highlighting their associations with various ACVD-related outcomes. For example, both the network regulator inter-alpha (globulin) inhibitor 3 (ITIH3) and the eigenprotein for that subnetwork were significantly associated with carotid plaque burden (Figure 4C). When considering the network regulators, seven CPN subnetworks showed significant links to all six ACVD-related traits (Figure 5A), while for the eigenproteins, only one CPN subnetwork (ITIH3) was associated with all six traits (Figure 5B).

**Figure 4.**
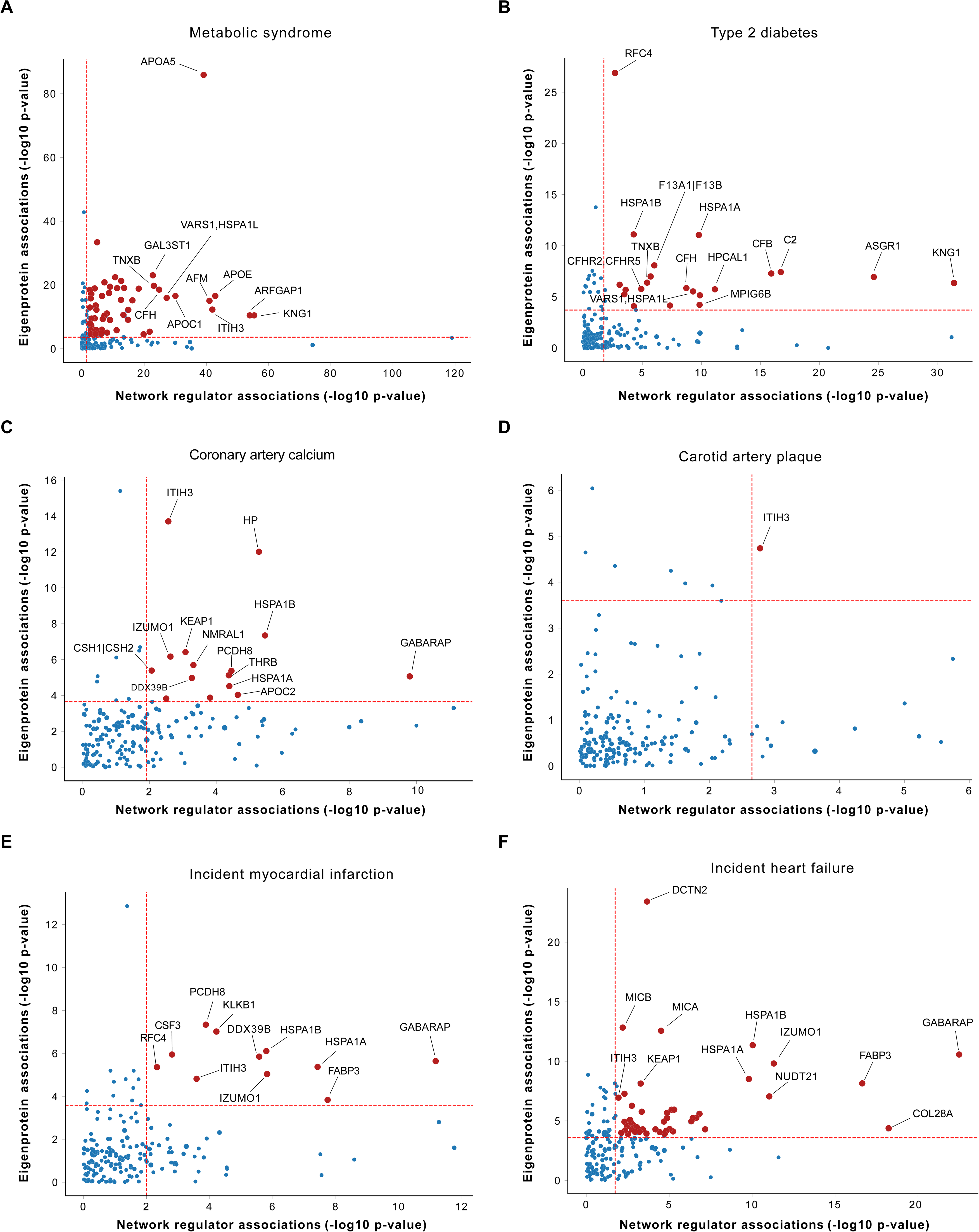
Correlation between network regulators and eigenproteins in relation to myocardial infarction and associated traits. The scatterplots (**A**-**F**) illustrate the correlation between the 182 network regulators and their corresponding network eigenproteins (1^st^ PC, explaining > 15% of the variance) in relation to myocardial infarction (MI) and cardiometabolic traits that contribute to atherosclerotic cardiovascular disease (ACVD), represented by -log10 P-values. Red dashed lines mark the 5% FDR thresholds for the respective axes.

**Figure 5.**
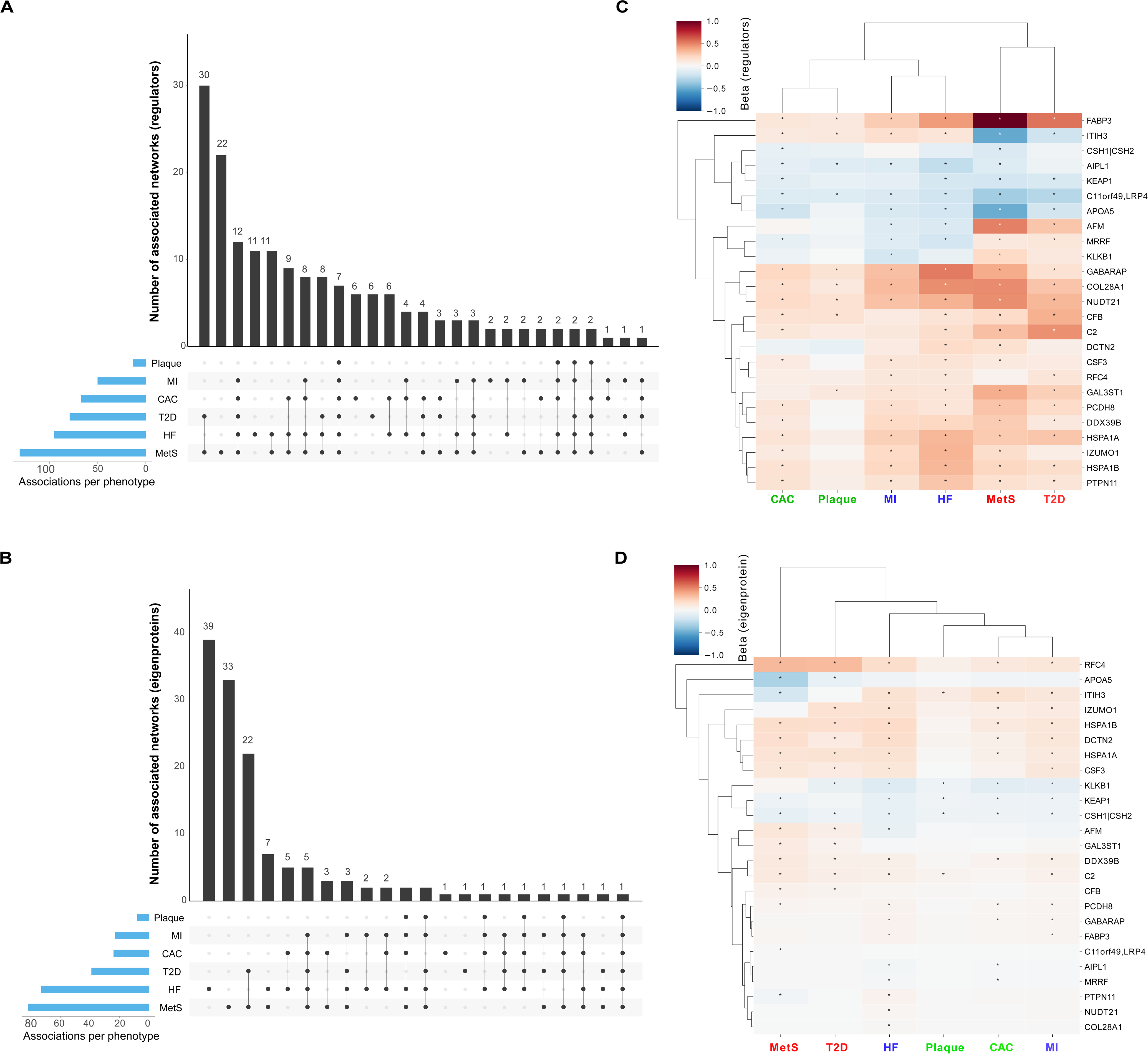
Intersection and clustering of subnetworks linked to myocardial infarction and related traits. The UpSet plots illustrate the intersections of the 182 causal protein networks (CPN) related to incident MI and related cardiometabolic trait associations with (**A**) the network regulators (FDR ≤ 5%) and (**B**) the CPN eigenproteins (FDR ≤ 5%). The heatmaps on the right depict the relationship between the top ranked subnetworks (**C**) network regulators and (**D**) eigenproteins and MI-related outcomes. Blue squares indicate a negative relationship between network regulators or eigenproteins and the outcome, while red squares represent a positive relationship. A star within a box denotes a significant association between proteins and outcome, based on either an FDR estimate of <5% (for network regulators) or a Bonferroni correction with adjusted P < 0.05 for the eigenproteins. <u>Abbreviations</u>: CAC, coronary artery calcium; Plaque, carotid artery plaque burden; T2D, type 2 diabetes (prevalent); MetS, metabolic syndrome; MI, incident myocardial infarction; HF, incident heart failure.

We ranked these subnetworks based on their associations with incident MI and other ACVD relevant outcomes, assigning equal weight to both the network regulator and its corresponding eigenprotein (Supplementary Tables S5 and S6). This approach identified 25 top-ranked subnetworks (arbitrary rank score ≥ 7; Table 2). Clustering the network regulators based on their associations with these outcomes revealed that the six core traits align with the three established etiological axes of MI (Figure 5C), as depicted in Figure 1. In contrast, the corresponding eigenprotein associations drive a distinct clustering of phenotypes, grouping CAC, carotid plaque burden, and MI together (Figure 5D), which represent the key outcomes associated with atherosclerosis^31^. The top ranked CPN subnetworks in Table 2 were largely retained when a stricter 30% eigenprotein variance threshold was applied (Supplementary Tables S7 and S8). Associations of all top ranked network regulators with incident MI and/or related traits are illustrated in Supplementary Figure S8. In summary, both the top ranked network regulators and their corresponding eigenproteins capture the established multi-dimensional axes of ACVD development, albeit through slightly different mechanisms.

**Table 2.**
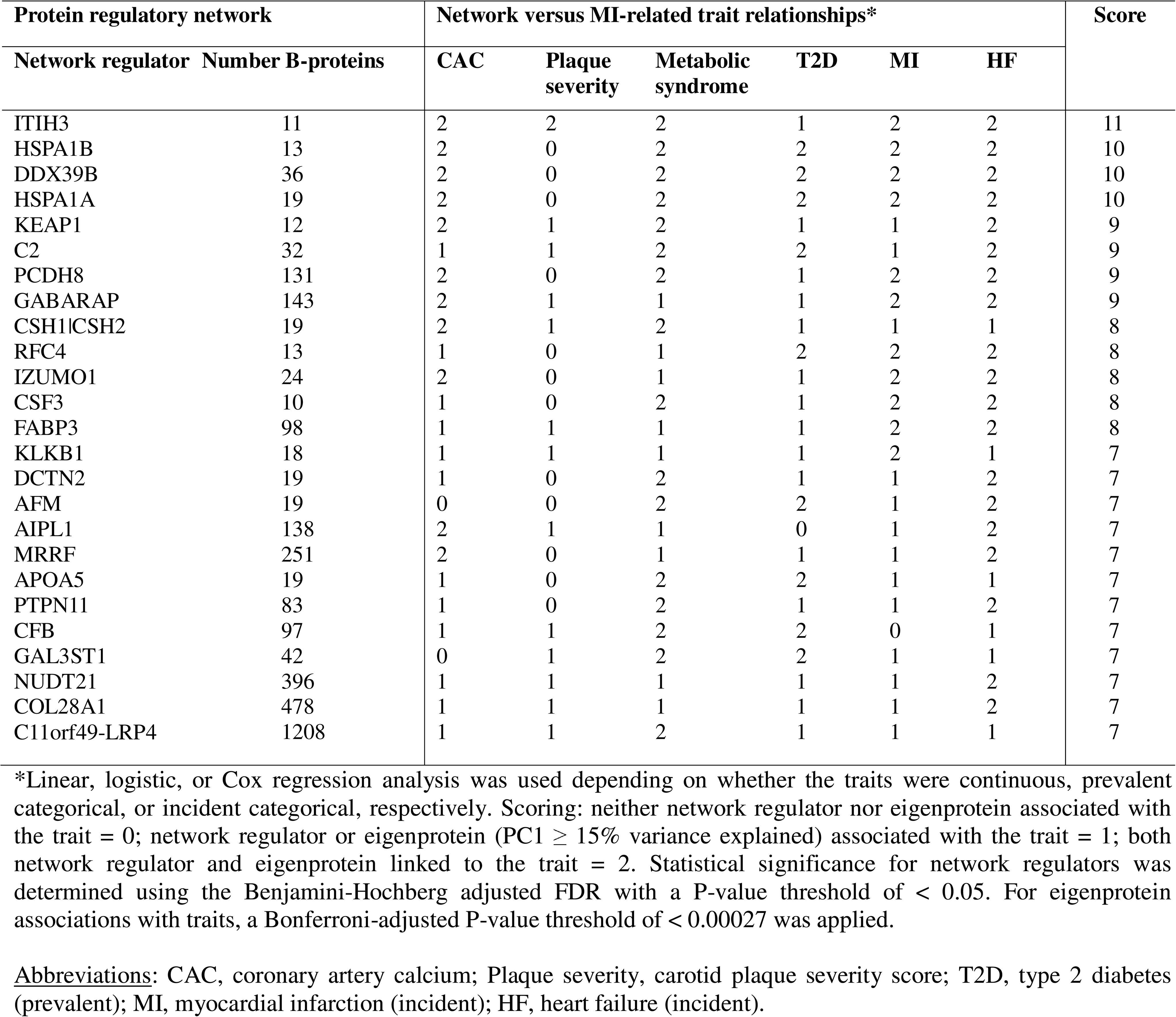
The top ranking (score ≥ 7) causal protein networks related to incident MI and associated traits.

### The top ranked subnetworks exhibit a strong degree of interconnectivity

Having identified subnetworks of the global CPN that are associated with different aspects of ACVD, we were interested in comparing structural similarities between these subnetworks. Among the 25 top ranked subnetworks (Table 2), we find examples of clusters of proteins that are co-regulated by several network regulators (Figure 6A). Furthermore, target similarity analysis identified clusters of CPN subnetworks that are both correlated through their eigenprotein and share similar targets (Figure 6B). Interestingly, we find a large group of negatively correlated subnetworks which shares similar targets, indicating co-regulation by distinct network regulators with opposing directional effects.

**Figure 6.**
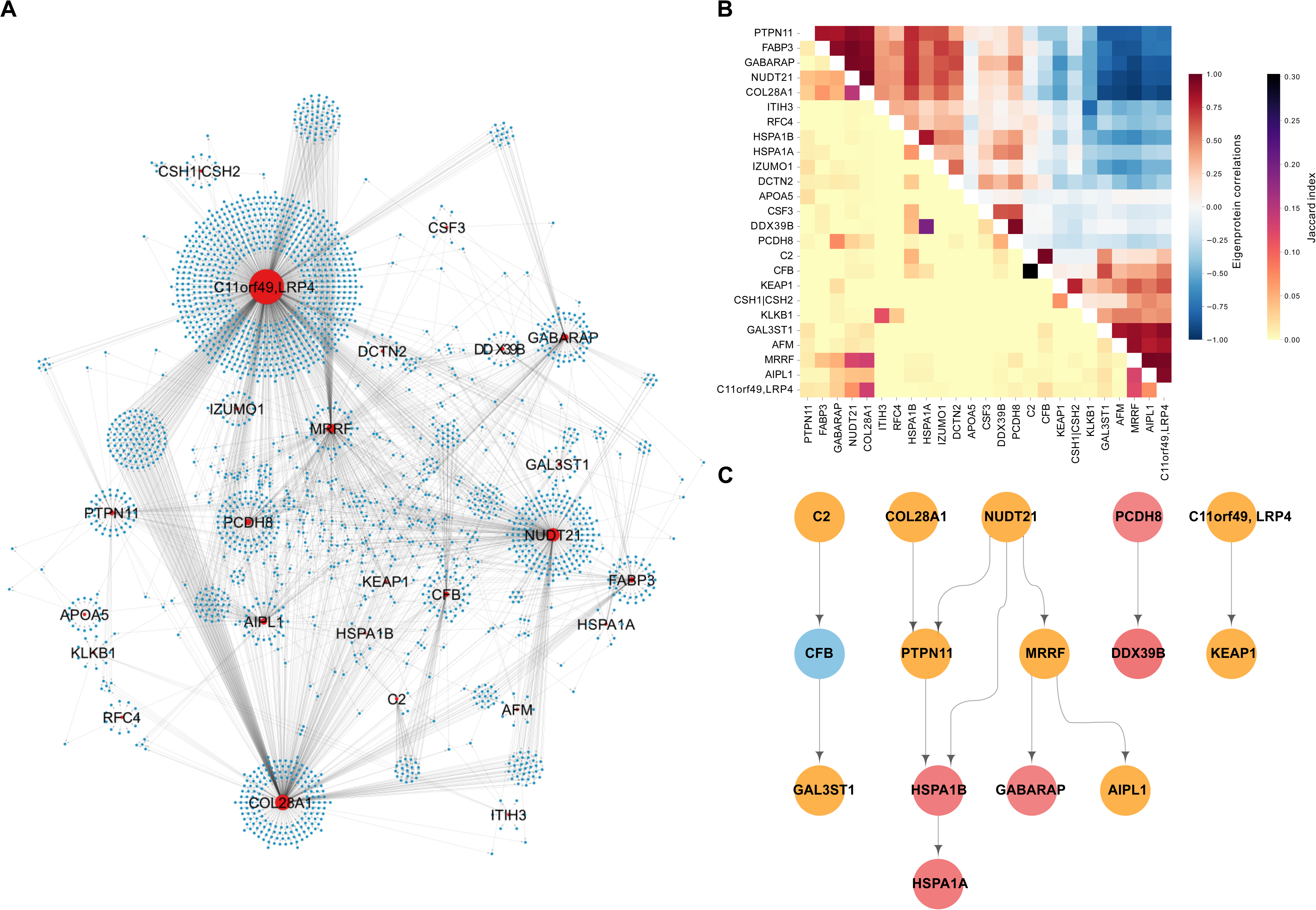
Interconnectivity between the top ranked causal protein networks associated with myocardial infarction and/or related traits. (**A**) Network visualization of the interconnections among the top ranked causal protein networks (CPN) for incident myocardial infarction (MI) and associated traits, where the variance explained by network eigenproteins is over 15% (see Table 2). Red nodes represent network regulators, while blue nodes denote target proteins. (**B**) Heatmap displaying the similarity between CPN subnetworks. The upper right triangle shows hierarchical clustering based on pairwise Pearson correlations between network eigenproteins. The clustering leaf order of the Pearson correlations has been applied to the lower left triangle, which represents pairwise Jaccard Index scores between the same networks. (**C**) Hierarchical representation of interacting network regulators where color indicates degree of association with incident MI. Blue = no association to incident MI, yellow = either eigenprotein or network regulator associated, red = both eigenprotein and network regulator are linked to incident MI.

Similar to the global CPN, we observed multiple levels of regulation among the network regulators of the top ranked networks (Figure 6C). The connected subnetworks form distinct modules that largely preserve this structure, even when the previously removed edges are reintroduced (Supplementary Table S1). This includes, for example, a cascade involving the network regulators C2, CFB, and GAL3ST1 (Figure 6C), as well as the CFHR1 network regulator when applying a more stringent eigenprotein variance threshold (Supplementary Figure S9A-B). This is noteworthy because CFHR1, C2 and CFB are all integral components of the same complement cascade, whereas GAL3ST1 is an enzyme involved in the biosynthesis of glycosaminoglycans, which play vital roles in various biological processes, including inflammation and vascular remodeling^32^. The role of the complement system in inflammatory processes and cardiac health is well-established, particularly in relation to myocardial tissue injury^33^. Another intriguing example involves a pathway of five proteins: COL28A1, NUDT21, PTPN11, HSPA1B, and HSPA1A (Figure 6C). Notably, all but COL28A1 are enriched in the hematopoiesis pathway (g:Profiler, P-value = 0.018), essential for blood cell formation and development from hematopoietic stem cells. This process is critical for maintaining homeostasis and has broad implications in atherosclerosis^34^. Interestingly, mutations in the mouse *Col28a1* gene are linked to abnormal blood vessel morphology, according to the MGI database^35^. These cascade-related proteins may collectively influence inflammation, vascular remodeling, atherosclerosis, and blood vessel integrity, supporting their observed associations with incident MI and related traits in this study.

Finally, comparing the top ranked CPN subnetworks with the previously published serum protein co-regulatory network from the AGES study^21^ revealed substantial overlap between protein clusters in both network types (Supplementary Table S9, Supplementary Figure S01A–D, Supplementary Text). This overlap is evident in two ways: first, many CPNs align with the same co-regulatory module; second, when a single CPN intersects with multiple co-regulatory modules, these modules often belong to the same super-cluster of correlated co-regulatory modules (Supplementary Text). Overall, a significant relationship exists between the circulating CPNs and the co-regulatory networks, despite fundamental differences in the methodologies used to reconstruct them.

### Additional links between the top ranked networks and ACVD

Several pathways were enriched among the network regulators and their targets from the top ranked subnetworks listed in Table 2 (Supplementary Table S10, Supplementary Figures S11 and S12). These included many pathways previously associated with ACVD pathophysiology (Supplementary Text). While most of the enriched pathways differed between the network regulators and target members, both sets shared enrichment in the cellular response to heat shock (Supplementary Text). We investigated known functional and physical protein-protein interactions among the top-ranked serum protein network regulators using the STRING PPI database^36^ and found significant enrichment in interaction levels (P = 0.047). This reflected some of the previously identified interactions shown in Figure 6A and provides additional insights, such as interactions involving the top ranked network regulator ITIH3, which has connections with KLKB1, APOA5, and AFM (Supplementary Figure S13).

We performed colocalization and two-sample MR analysis on the network regulators listed in Table 2 to further investigate potential causal relationships with ACVD related outcomes (see Methods). This analysis identified the network regulators APOA5, DDX39B, HSPA1A, LRP4/C11orf49, with significant (FDR < 0.05) causal relationship with MI and/or related traits based on findings from both the MR and colocalization analyses (Supplementary Tables S11 and S12). Variants in the gene encoding APOA5 have previously been linked to coronary atherosclerosis and coronary revascularization^37^, as well as hyperlipoproteinemia in several studies^38^. In the current study, APOA5 showed a strong causal relationship with both MI and MetS, supported by findings from both colocalization and MR analyses (Supplementary Table S12). The gene encoding HSPA1A has been causally associated with T2D and its microvascular complications through both MR and colocalization analyses^39^, while our findings establish a causal relationship between HSPA1A and MetS (Supplementary Table S12). Finally, the locus containing the genes encoding C11orf49 and LRP4 has been linked to cardiomyopathy^37^ and venous thromboembolism^40^, respectively. C11orf49 and LRP4 were each found to have a causal relationship with MI in the AGES study (Supplementary Table S12), though this was mediated through a shared instrument. These findings provide further evidence for the causal role of these network regulators, and likely their corresponding targets, in the development of ACVD.

Several other top ranked network regulators have been previously linked to ACVD and/or cardiometabolic traits that contribute to the disease (Supplementary Text), including a functional genetic variant in the gene encoding the network regulator ITIH3, which has been associated with MI^41^. This top ranked regulator is expressed in vascular smooth muscle cells and macrophages within human atherosclerotic lesions^41^. Interestingly the data-driven reconstruction of the CPN reveals that the roots of the global network regulating a diverse array of network targets converge on ITIH3 at the base of the network (Figure 3), suggesting its involvement in many regulatory pathways. Other notable associations include genetic variants in the *KEAP1*, *C2*, *GABARAP*, *KLKB1*, *PTPN11*, and *CFB* genes, which have been linked to various ACVD related outcomes (Supplementary Text). Thus, for nearly half of the top-ranked network regulators, this study, along with others, reveals additional specific connections to ACVD-related traits, beyond those identified through the workflow of the current study.

## Discussion

Atherosclerotic cardiovascular disease remains the leading cause of death worldwide^1^, emphasizing the urgent need for early detection and innovative preventive strategies. Clinical complications to ACVD like MI, stroke, and its longer term complication HF, arise from a complex interaction of both local (cardiovascular) and systemic (non-cardiovascular) factors, highlighting the importance of a broad systems-level approach to understand their intricate etiology. To address this, we employed a causal inference approach to reconstruct the first large-scale circulating CPN in humans. We identified 185 CPN subnetworks, many of which were strongly associated with the onset of MI and/or HF, as well as with upstream clinical risk factors essential to ACVD development. The strongest CPN subnetworks associations clustered these phenotypes according to their localization within the key organ axes central to ACVD related pathophysiology. This study begins to uncover the underlying systemic mechanisms, reflected in serum proteins, that drive processes leading up to the onset of MI and its long-term consequence, HF.

The central dogma of molecular biology outlines the process of information transfer from DNA to proteins. Proteins have the capability to influence phenotypes, including disease, through biological networks. The capacity to acquire measurements of thousands of proteins from a single blood sample has opened new paths for monitoring health and disease in greater depth than ever before^24,42^. We identified the first circulating protein co-regulatory networks in humans, linking it to the genome in an unbiased manner and elaborating on those findings to highlight its connections to complex disease^21^. Despite being highly informative, the previously described co-regulatory network^21^ is undirected, which means that the causal relationship between nodes is unknown. More recently, large-scale directed single-tissue and cross-tissue gene-regulatory co-expression networks have been reconstructed across multiple tissues related to CAD, providing a comprehensive mechanistic framework for understanding the etiology of the disease^9^. Causal inference and causal networks CPN present an ideal framework for graphically modeling complex systems because it allows for the transmission of prior information about a system and the formulation of concrete hypotheses for follow-up research^43^. As such, causal inference is situated between purely data-driven machine learning and detailed mechanistic modelling approaches. Causal network inference, however, is a computationally demanding and complex process, often restricted to small-scale systems. To overcome this, we developed a new approach that outperforms previous methods in both efficiency and coverage, and proving particularly effective when both genotype and molecular node data are available for the same sample^17,18^. In this study, we used this approach within the single-center population-based AGES study. Each participant provided essential genotype, proteomic, and clinical data, which allowed for the identification of the circulating CPN at a low FDR threshold, as well as the detection of subnetworks associated with the future onset of ACVD.

Although circulating co-regulatory networks and CPNs are reconstructed using different methodologies, we observed a significant overlap between these two types of networks. CPNs comprise a single regulator and its target protein nodes, while co-regulatory networks encompass a much broader scope. In fact, because many CPNs interact and often overlap within the same co-regulatory module, they may converge to create a larger co-regulatory module. The significant overlap between the two types of networks is not entirely unexpected, as co-regulatory serum protein networks have been shown to be strongly influenced by genetic factors^21^, and proximal pQTL were employed as instrumental variables in reconstructing the CPN. This suggests that the overlapping protein clusters shared between the two network types are likely influenced by the same genetic factors. However, CPNs offer additional insights for causal relationships between proteins that are not evident in co-regulatory networks. In other words, the CPN can illustrate how changes in one protein node can regulate another node even in the presence of unobserved confounding factors, effectively distinguishing between the effects of correlation and causation. This ultimately improves the modeling of complex disease etiology.

A key strength of our study is the high-quality data from a large-scale, prospective, population-based cohort, which includes detailed phenotype information for each participant, extensive coverage of circulating proteins, and comprehensive genomics data. This study, however, has several limitations that must be acknowledged. The present findings are limited to serum proteins and may not fully capture the MI-related pathobiology in solid tissues, such as the arterial wall and heart. Moreover, the study does not encompass the entire serum proteome, which is still being mapped; however, it remains one of the largest studies of its kind to date. Regarding the CPNs, we still do not have a clear understanding of the nature of the edges in our serum protein networks, especially whether they represent direct or indirect regulation mediated by processes within or across tissue boundaries. Our findings, however, indicate that while the CPNs partially reflect protein-protein interactions, they may also capture interactions between nodes across tissues. Additionally, a high number of indirect interactions have been observed in these networks; however, it is not currently possible to determine which of these are true instances of FFLs, commonly enriched in biological networks, or false positives. Developing causal methods that use higher-order mediation tests between triplets of proteins could address this issue. Current mediation tests, however, suffer from hidden confounding, which is accounted for within randomization-based approaches. Finally, it should be noted that validating our CPNs is challenging because complimentary external data is lacking, unlike tissue-specific GRNs based on gene expression QTLs, which have some external transcription factor to target data available^44^. Therefore, the extent to which these edges can be independently validated remains to be determined.

Among complex traits, ACVD and its associated clinical complications, including MI, stroke, and HF, represent the highest level of complexity and continue to be a leading cause of morbidity and mortality in industrialized nations. ACVD arises from the complex interplay of cardiometabolic disorders, which collectively drive the progression of coronary plaques over decades. These cardiometabolic factors, affecting the arterial wall, involve multiple tissues and are shaped by intricate genetic and environmental influences. While this high level view of the disease has been recognized for years, the detailed endocrine signaling linking the tissues and biological processes involved remain poorly understood. The extensive measurement of over 7,500 proteins has facilitated exploration of the architecture of the serum proteome and the regulatory relationships of blood proteins, many of which were associated with ACVD, with some demonstrating causal links to the disease. These regulatory relationships were deeply interconnected, forming cascades of protein nodes or networks that elucidate the directionality and collective mechanisms driving ACVD. The serum protein networks span tissue boundaries, linking key tissues and organs involved in the etiology of ACVD. This work begins to reveal, at the molecular level, the cross-tissue coordination or systemic homeostasis required for disease manifestation. The initial characterization of this network lays the foundation for formulating hypotheses and directing future research on the etiology of ACVD.

## Material and Methods

### Study population

Cohort participants aged 66 through 96 years at the time of blood collection were from the AGES^25^, a single-center, prospective, population-based study of older adults (N = 5,764, mean age 76.6±6 years). AGES was formed between 2002 and 2006, and its participants were randomly selected from the surviving members of the established 40-year-long population-based Reykjavik study^45,46^. The Reykjavik study, a prospective cardiovascular survey, recruited a random sample of 30,795 adults born between 1907 and 1935 who lived in the greater Reykjavik area in 1967, that were examined in six phases from 1967 to 1996^45,46^. The AGES measurements, which include for instance brain and vascular imaging, are designed to assess four biologic systems: vascular, neurocognitive (including sensory), musculoskeletal, and body composition/metabolism^25^. All AGES participants are of European ancestry. The AGES study was approved by the National Bioethics Committee in Iceland that acts as the institutional review board for the Icelandic Heart Association (approval number VSN-00-063, in accordance with the Helsinki Declaration) and by the US National Institutes of Health, National Institute on Aging Intramural Institutional Review Board.

The study comprised MI patients who met the MONICA criteria for definite MI as previously described^45^. The criteria for HF were based on clinical symptoms and signs, chest X-rays, and, in many cases, echocardiographic findings from hospital records, which were adjudicated by examining every record for both prevalent HF, i.e., had HF at the baseline visit, and incident HF, i.e., HF diagnosed after the baseline visit. The incident HF cases were free of HF diagnosis at the baseline visit, but who were later hospitalized and diagnosed (hospital discharge ICD-10 diagnosis codes starting with I50) with HF during the follow-up period of eight years. Each patient’s thorough medical records were subsequently adjudicated by a cardiologist to confirm the diagnosis of symptomatic HF, and the date of the incident HF event documented. Among the criteria were symptoms such as shortness of breath that could be ambulatory, and signs of pulmonary edema. Type two diabetes (T2D) was determined from self-reported diabetes, diabetes medication use, or fasting plasma glucose ≥ 7 mmol/L according to American Diabetes Association guidelines.^47^ Metabolic syndrome (MetS) was defined by three or more of the following: Fasting glucose ≥ 5.6mmol/L, blood pressure ≥ 140/90, triglycerides ≥ 1.7 mmol/L, high-density lipoprotein (HDL) cholesterol <0.9 mmol/L for males or <1.0 mmol/L for females, body mass index (BMI) > 30kg/m^2^. Systolic and diastolic blood pressure were measured twice with subjects in a supine position using a Mercury sphygmomanometer. Lipoproteins and plasma glucose levels were measured on fasting blood samples. Triglycerides (TG) were measured using enzymatic colorimetry (Roche Triglyceride Assay Kit), HDL cholesterol with an enzymatic in vitro assay (Roche Direct HDL Cholesterol Assay Kit), and glucose was measured using photometry (Roche Hitachi 717 Photometric Analysis System). Coronary artery calcium (CAC) was quantified using the Agatston scoring method^48^, which was reviewed independently by four image analysts. Phantom-adjusted CAC was expressed as a sum score for all four coronary arteries, as previously described in greater detail^49^. The use of ultrasound imaging to assess the presence and severity of carotid plaque in the AGES population has been detailed elsewhere^50,51^.

### Proteomics profiling

Blood samples were collected at the AGES-Reykjavik baseline visit after an overnight fast, and serum samples prepared using a standardized protocol and stored in 0.5mL aliquots at -80°C. Serum samples collected from the inception period of AGES, i.e., from 2002 to 2006, were used to generate proteomics data. Before the protein measurements were performed, all serum samples from this period went through their first freeze-thaw cycle. Protein levels in serum from 5,376 individuals of the AGES were determined using a multiplex SOMAscan proteomic profiling platform which employs aptamers or Slow-Off rate Modified Aptamers (SOMAmers) that bind to target proteins with high affinity and specificity. Here, 7,523 aptamers mapping to 6,586 UniProt IDs were measured in total of 8,592 samples (two time points) using the most recent SomaScan_v4.1 platform^52^. The aptamer-based platform measures proteins with femtomole (fM) detection limits and a broad detection range (>8-log dynamic range) of concentration^53^. To prevent biases related to batch or processing time, the order of sample collection and separate sample processing for protein measurements were randomized, and all samples run as a single set at SomaLogic Inc. (Boulder, CO, US). All aptamers that passed quality control had median intra-assay and inter-assay coefficient of variation, CV < 5%. Hybridization controls were used to correct systematic variability in detection and calibrator samples of three dilution sets (20% (1:5), 0.5% (1:200), and 0.005% (1:20,000)) were included so that the degree of fluorescence was a quantitative reflection of protein concentration. The adaptive normalization by maximum likelihood (ANML) method was employed to normalize Quality Control (QC) replicates and samples using point and variance estimations from a healthy external reference population (n = 1000). Consistent target specificity of aptamers was indicated by direct (through mass spectrometry) and/or indirect validation^21^.

Some proteins were targeted by more than one aptamer. In such cases, individual aptamers had distinct binding sites (epitopes) or binding affinity^21^. The single gene *NPPB*, for example, produces three protein products: full-length BNP, NT-proBNP, and BNP32, each of which are targeted by different aptamers. Duplicate aptamers to single pass transmembrane proteins (one to extracellular domain and another to intracellular loop), aptamers targeting multimers (e.g., interleukins), and duplicate aptamers generated in distinct expression systems are further examples. Finally, 233 aptamers were derived from mouse-human protein chimeras (used as SELEX input) to target proteins from both species.

Before the analyses, protein data were centered, scaled and Box-Cox transformed,^54^ and extreme outlier values were excluded, defined as values above the 99.5^th^ percentile of the distribution of 99^th^ percentile cutoffs across all proteins. Prior to reconstruction of the causal protein networks, the data were adjusted using a linear model to account for age and sex.

### Genotype data and the detection of cis-acting variants

The genotype data includes assayed and imputed genotype data for 5,636 AGES participants^55^. The genotyping arrays used were Illumina Hu370CNV and Illumina GSA BeadChip, which were quality controlled by eliminating variants with call rates <95% and Hardy Weinberg Equilibrium (HWE) P-value < 1 × 10^−6^. The arrays were imputed against the Haplotype Reference Consortium imputation panel r1.1 and post-imputation quality control was performed separately for each platform. Variants with imputation quality *r*^2^ < 0.7, minor allele frequency < 0.01, as well as monomorphic and multiallelic variants, were removed before merging the platforms to generate a dataset with 7,506,463 variants for 5,656 AGES individuals as previously described^55^. These variants were associated to each of the aptamers on the v4.1-7k serum protein panel to identify proximal (*cis*-acting) pQTLs, in the same way as previously described^55^. We applied a 300kb genomic window spanning each protein expressing gene in the v4.1-7k serum protein panel to map out *cis*-acting pQTLs after accounting for the number of single nucleotide polymorphisms (SNPs) in each window. We then corrected for multiple testing using the Storey-Tibshirani procedure for q-value estimation^56^. The *cis*-acting pQTLs serve as instrumental variables for reconstruction of the causal serum protein networks, which are described below.

### Reconstruction of the circulating causal protein network

A circulating Causal Protein Network (CPN) was reconstructed from causal relationships between serum proteins derived using a Mendelian Randomization (MR) framework^57^. In this model, causal relationships between protein pairs are estimated between an exposure protein (*A*) and outcome protein (*B*), using a *cis*-acting pQTL for *A* as a causal instrument. We selected all proteins which had at least one valid *cis*-acting pQTL as our A-proteins at FDR ≤ 5%, after correcting for multiple testing using the Storey-Tibshirani procedure^56^. In each pairwise causal test, the lead protein regulatory SNP (pSNP) for *A* (lowest P-value) was selected as *E* to be used as an instrumental variable in accordance with the core instrumental variable assumptions^57,58^: 1) the instrumental variable should be strongly associated with the exposure (*A*), 2) the instrumental variable should only be associated with the outcome (*B*) through the exposure (*A*), and 3) the exposure (*A*) should not be associated with any potential confounders.

Causal estimates between proteins were measured using the tool Findr^18^ (version 1.0.8) in Python (version 3.10.6) using individual level protein expression levels for *A* and *B* and genotypes for *E*. Before inferring casual estimates, the data were transformed using a rank-based inverse normal transformation within the Findr package. The product of the secondary linkage test (P2) and controlled test (P5) from Findr^18^ was used to estimate the posterior probability (PP) of P(*A*→*B*). The secondary linkage test measures the PP of association between *E* and B and the controlled test assesses the PP of the dependence between *A* and *B* following adjustment with E to exclude that E has independent effects on A and B^59^. We estimated P(*A*→*B*) for all A-proteins with a valid instrumental variable, where *B* was every other protein in our dataset. We estimated a global FDR as 1 minus the mean of all PP for P(*A*→*B*) and then filtered PP P(*A*→*B*) to achieve the desired FDR threshold, as shown previously^60^. Networks were reconstructed from causal interactions that fell below this FDR threshold, where parent nodes were *A* and child nodes were *B*, with edges represented as P(*A*→*B*). In instances where there were multiple aptamers targeting the same protein, we selected the *A* proteins with the largest number of targets as the representative aptamer for this protein. When examining hierarchy between regulators, we converted edges between *A* proteins to a directed acyclic graph (DAG) using a greedy heuristic as implemented in Findr^18^, which selects the most significant edges in an iterative fashion to avoid cycles. This is done to identify any hierarchical structure between A-proteins, and once a DAG has been constructed the removed edges can be reintroduced to examine how well any hierarchical structure is preserved within the complete networks. Network visualization was performed using Cytoscape (version 3.10.1).

As pleiotropy is a concern in MR analyses, we took steps to account for instances where two or more exposure proteins either shared or had instrumental variables in high linkage disequilibrium (LD)^58^. We assigned proteins to the same LD block if they shared the same lead pSNP, or if they had different pSNPs in medium or high LD (*r*^2^ ≥ 0.5). For all *A* proteins within an LD block, we calculated the intersection of targets as a ratio of the union of targets (*I*) in a pairwise manner between target sets. We resolved *A* proteins as independent networks when *I* < 0.6, in cases where *I* > 0.6 we collapsed the union of all targets within a single unresolved network. In cases where two or more *A* proteins were mutual targets of each other, we considered these networks as unresolved. Where there were more than two A-proteins in a single LD block, an A-protein must have I < 0.6 across all other A-proteins to be considered independent. We did not encounter any instances of A-proteins that were shared between separate LD blocks.

We defined protein subnetworks as a single A-protein and all targets of that A-protein, i.e. a single regulator and its targets. The average expression profile of a subnetwork was estimated as an eigenvector of all proteins in a subnetwork across all samples, described here as an eigenprotein^61^. We calculated the first principal component (PC) from the expression profiles of all proteins within a subnetwork using principal component analysis (PCA), via the PCA function from the Python library scikit-learn^62^ (version 1.1.2). If PC1 explained > 15% of the variance of the subnetwork, we used this principal component as an eigenprotein for this subnetwork in downstream analyses, and for subnetworks where the variance was < 15%, PC1 was not taken forward. Three CPN subnetworks did not meet this criterium and therefore did not have a valid eigenprotein.

### Network robustness analyses

To examine the impact of varying sample size on network inference, we randomly sampled subsets of individuals in different sizes for the reconstruction of networks. At each sample size threshold (n = 500, 1000, 2000, 3000, 4000, 5000), we randomly sampled three subsets from the pre-processed protein expression data and reconstructed a network for each subset using the previously outlined approach. We termed the primary network, which was reconstructed using all samples, as the ground-truth network. The ground-truth network was represented as a flattened binary matrix (A,B), where 1 was indicative of the presence of an edge between A→B, and 0 for absence of an edge. Each sub-sampled network was represented as a similar flattened matrix (A,B), but where the values were populated by the PP of P(A→B). We tested how well each sub-sampled network captured the structure of the ground-truth network by calculating the Receiver Operating Characteristic Area Under the Curve (ROC AUC) for each sub-sampled network against the ground-truth network using the roc_auc_score function from scikit-learn (version 1.1.2)^62^. We then calculated the mean AUC of the repeated sub-sampled networks at each sample size threshold. We also calculated Precision-Recall for each sub-sampled network against the ground-truth network using the precision_recall_curve function, also from scikit-learn^62^.

### Variance explained model

To identify independent *cis*-acting protein SNPs, a ±150kb window was specified around each lead variant to define the region of interest for association testing. Using the GCTA software (version v1.94.1)^63^, the cojo-slct parameter was applied using a forward model selection approach. The default collinearity cutoff of 0.9 was used and the P-value threshold was set at 0.00763, which corresponds to the highest P-value maintaining an FDR below 5%. Having identified independent *cis*-acting protein SNPs for every network regulator, we estimated the proportion of variance in protein expression that could be explained by *cis*-acting pQTLs using multiple linear regression. For each protein we fitted a linear model in R (version 4.3.2), where the genotypes for the independent *cis*-acting SNPs act as the explanatory variables for protein expression. The adjusted coefficient of determination (adjusted *r*^2^) from this model was used as an estimate of the variance explained by the *cis* component for each protein, and the mean adjusted *r*² was then calculated across all network regulators.

We also estimated the variance in target protein expression that could be explained by *cis-*acting pQTLs from the target regulators. More specifically, the influence of *cis* signals on network regulators (parental nodes) was assessed in relation to the expression of 5,459 target proteins within the CPN, including 162 target proteins that also served as regulators for other proteins. For every target protein, we fitted a linear model, as described previously, using the genotypes of all independent *cis*-pQTLs for the regulators (parental nodes), in addition to any *cis*-pQTLs for the target itself. We then calculated the difference in variance explained by local and parental *cis*-pQTLs combined, to that explained by *cis*-acting genetic variation alone. If the target protein did not have any *cis*-pQTLs, then the variance explained by the *cis* component was set to 0. Any unresolved networks were excluded from this analysis.

### Comparison with a reference database of protein-protein interactions

We used the human integrated protein-protein interaction reference database (HIPPIE) version 2.3^30^ to identify experimentally derived protein-protein interactions (PPIs) that have been captured by the serum CPN described in this study. We accessed all 289,112 PPIs from the HIPPIE database, which have been scored as a weighted sum, based on the reliability of the evidence underpinning the interactions and the number of studies detecting each interaction. There are 262,346 PPIs scored at medium confidence (confidence score > 0.63) and 77,630 scored at high confidence (confidence score > 0.72), based on thresholds defined by HIPPIE authors. We then calculated the overlap between edges in the CPN with PPIs from HIPPIE at the different confidence thresholds. After which, we compared the number of common interactions between HIPPIE and the CPN to common interactions between HIPPIE and the edges from random networks. These networks were generated by randomly sampling proteins from the complete set of measured AGES proteins, to produce the same number of edges as in the CPN. This process was performed 10 times and the mean number of edges captured, in addition to the standard deviation, was calculated at each of the different confidence thresholds. We then calculated a z-score comparing the number of edges captured from the true vs random networks at each confidence threshold as: z = (observed overlap - mean random overlap) / stdev random overlap. This was then converted to a p-value as: p-value = 1 – cdf(z-score), using the norm.cdf() function from Scipy Stats^64^.

### Network topology analyses

We identified weakly connected components (i.e., groups of nodes in the CPN where every node can be reached from any other node, regardless of the direction of the edges) in the complete CPN and after removal of the regulators with the most targets applying the weakly_connected_components function from the Graphs.jl (v1.12.0) package using Julia v1.11.1. We obtained a hierarchical layout of the A-protein DAG by first defining root nodes as A-proteins without incoming edges. We then defined the level of each A-protein as the shortest distance (number of edges in a shortest path) from a root node to the protein of interest. Shortest paths were computed using the dijkstra_shortest_path function from the Graphs.jl (v1.12.0) package using Julia v1.11.1. To further simplify the structure of the DAG, we performed transitive reduction using the “transitive reduction” function from the Graphs.jl (v1.12.0) package from Julia. Transitive reduction reduces the original DAG G to a new DAG G’ with the fewest possible edges such that, if there is a path from a vertex x to a vertex y in G, there must also be a path from x to y in G’, and vice versa.

### Colocalization and Mendelian randomization analysis

We used colocalization analysis to identify *cis*-pQTLs that shared a common signal with six different phenotypic traits using the R package coloc (version 5.2.3)^65^ and publicly available GWAS summary statistics from individuals with European ancestry (Supplementary Table S10). We first identified all proteins that had a *cis-*pQTLs (FDR < 5%) that shared at least 1 common *cis*-SNP with the GWAS trait (P < 5×10^-8^) and extracted the summary statistics for all SNPs within ±150 Kb of the transcription start site for the cognate encoding gene. We then estimated the probability of a shared causal variant (PP.H4) using approximate Bayes Factor colocalization *via* the coloc.abf function using a threshold of PP > 0.9.

The causality of selected network regulators was assessed using a two-sample *cis*-Mendelian randomization (MR) approach^66^. Genetic instruments (*cis*-acting pSNPs) were identified within ±500 kb of the protein-coding region based on internal independent study data, filtered for statistical significance (P < 0.05/number of *cis*-region SNPs), and matched to external GWAS outcomes. These SNPs were clumped for LD (r² ≥ 0.2) using PLINK v1.9^67^, and AGES genotype data. MR estimates were calculated using the Wald ratio for single-SNP associations and GWLS for multi-SNP analyses as previously described^68^. Significant results (FDR < 0.05) were deemed causal candidates.

### Statistical analysis and functional enrichment analysis

The relationship between serum protein (and eigenprotein) levels and quantitative phenotypes was evaluated using linear regression controlling for age and sex, whilst the relationship between serum protein (and eigenprotein) and prevalent disease was examined cross-sectionally using logistic regression with age and sex adjustments. The associations between serum proteins (and eigenprotein) and incident disease were evaluated longitudinally using the Cox proportional-hazards model^69^, with a median follow-up period of 5.35 years [2.59, 8.16] years for incident MI. Associations with Benjamini-Hochberg FDR < 0.05 were considered statistically significant. To identify enriched gene ontology (GO) terms and pathways within network targets, we carried out formal function enrichment analysis using gprofiler2 (version 0.2.2), the R interface of g:Profiler (R version 4.3.2). We used a custom background of all measured proteins in the AGES aptamer-based assay and corrected for multiple testing using Benjamini-Hochberg correction with a cut-off of FDR < 0.05 to identify any enriched terms across the following categories; GO:MF, GO:BP, GO:CC, KEGG, REACTOME, Wikipathways, miRTarBase, Human Protein Atlas and Human phenotype ontology.

## Acknowledgement

The authors acknowledge the contribution of the Icelandic Heart Association (IHA) staff to the AGES-RS, as well as the involvement of all study participants. The proteomics work was carried out in collaboration with Novartis Biomedical Research (NIBR). National Institute on Aging (NIA) contracts N01-AG-12100 and HHSN271201200022C for V. G. financed the study. V. G. received funding from the NIA (1R01AG065596-01A1), and IHA received a grant from the Icelandic Parliament. T.M. acknowledges support from the Research Council of Norway (project number 312045), the European Union’s Horizon Europe (European Innovation Council) programme (grant agreement number 101115381), and the L. Meltzers Høyskolefond.

## Resource Availability

Data from the AGES Reykjavik study are available through collaboration under a data usage agreement with the IHA. All access to data is controlled via the use of subject-signed informed consent authorization. The time it takes to respond to requests varies depending on their nature and circumstances of the request, but it will not exceed 14 working days. All data supporting the conclusions of the paper are presented in the main text and freely available through supplementary data to this manuscript. All code used in this study is accessible at the following repository: https://github.com/sbankier/AGES_causal_protein_networks.

## Author contributions

S.B and V.E co-wrote the manuscript. S.B., V.E. and T.L.R.M. produced visualizations, and contributed to the conception and design of this research. S.B., V.E., T.L.R.M., Va.G., H.B. and T.J. performed the analyses. S.B., Va.G., and T.J. were responsible for data curation. All authors participated in reviewing, data interpretation and editing of the manuscript. All coauthors have approved the submitted version of the paper.

