## Supplementary Material for "Circulating causal protein networks linked to future risk of myocardial infarction"

### Supplementary Text

#### *Variance explained by parent nodes*

Having identified independent *cis*-acting protein SNPs for every network regulator, we estimated the proportion of variance in protein expression that could be explained by *cis*-acting pQTLs using multiple linear regression. For each protein we fitted a linear model in R (version 4.3.2), where the genotypes for the independent *cis*-acting SNPs act as the explanatory variables for protein expression. The adjusted coefficient of determination (adjusted $r^2$) from this model was used as an estimate of the variance explained by the *cis* component for each protein, and the mean adjusted $r^2$ was then calculated across all network regulators.

We also estimated the variance in target protein expression that could be explained by *cis*-acting pQTLs from the target regulators. For every target protein, we fitted a linear model, as described previously, using the genotypes of all independent *cis*-pQTLs for the regulators (parental nodes), in addition to any *cis*-pQTLs for the target itself. We then calculated the difference in variance explained by local and parental *cis*-pQTLs combined, to that explained by *cis*-acting genetic variation alone. If the target protein did not have any *cis*-pQTLs, then the variance explained by the *cis* component was set to 0.  Any unresolved networks were excluded from this analysis.

We examined the impact that *cis*-pQTLs for network regulators (parental nodes) had upon the expression of 5,459 target proteins in the CPN, including 162 target proteins that were also regulators for other proteins. For each target protein, we estimated the proportion of variance in protein expression (adjusted $r^2$) that could be explained by *cis*-pQTLs for all regulatory proteins, in addition to any independent *cis*-pQTLs for this protein. We found that the number of regulators a target protein has is correlated with how much variance can be explained by those regulators (Spearman R = 0.78) (Supplementary Figure S3A-B). We observed 32 cases where more than 50% of the variance in the expression of the target protein is explained by the cis-pQTLs of the parents alone, with no local *cis*-component contributing. Among these, 11 have less than 10 regulators. For example, DAP has 3 regulators: TXNRD3, KNG1 and HRG. KNG1 and HRG are within 1 KB on of each other on chromosome 3, and share 5 targets (20 targets each), however interestingly their instruments are not in LD ($R^2 < 0.2$). TXNRD3 is also on chromosome 3, but ~60 MB upstream. However, DAP is on chromosome 5 and together these *cis*-SNPs explain 82% of the variance in DAP protein expression, with no *cis*-acting signal for DAP being observed.

#### *Comparing the CPN with protein-protein interaction networks*

We used the human integrated protein-protein interaction reference database (HIPPIE)[1] to identify experimentally derived protein-protein interactions (PPIs) that have been captured by the serum CPN described in this study. We accessed all 289,112 PPIs from the HIPPIE database, which have been scored as a weighted sum, based on the reliability of the evidence underpinning the interactions and the number of studies detecting each interaction. There are 262,346 PPIs scored at medium confidence (confidence score > 0.63) and 77,630 scored at high confidence (confidence score > 0.72), based on thresholds defined by HIPPIE authors. We then calculated the overlap between edges in the CPN with PPIs from HIPPIE at the different confidence thresholds. After which, we compared the number of common interactions between HIPPIE and the CPN to common interactions between HIPPIE and the edges from random networks. These networks were generated by randomly sampling proteins from the complete set of measured AGES proteins, to produce the same number of edges as in the CPN. This process was performed 10 times and the mean number of edges captured, in addition to the standard deviation, was calculated at each of the different confidence thresholds.

We identified edges in the serum protein network (FDR = 1%) prior to LD resolution, that had previously been identified as PPIs using the HIPPIE database. PPIs from HIPPIE have been scored based on the strength of the supporting experimental evidence and the number of studies where and interaction has been detected. We identified 506 CPN edges that were also PPIs in

HIPPIE at any score, 429 at medium confidence and 216 at high confidence, using confidence thresholds that have been defined by the HIPPIE authors (Supplementary Figure S7). We then compared the overlap between HIPPIE and the CPN edges to the mean number of edges captured by networks generated through random sampling (Methods). We found that across all confidence thresholds, the true CPN consistently captured more edges than the mean captured by random networks: 332 edges at any score ($z = 14.6$, P-value $< 0.001$), 296 at medium confidence ($z = 10.3$, P-value $< 0.001$), and 94 at high confidence (z-score = 24.3, P-value $<0.001$) (Methods, Supplementary Figure S7). It should be noted that HIPPIE measures direct PPIs and does not account for indirect interaction between proteins, potentially mediated through tissue signaling which are captured by the CPN. Therefore, it is only possible to validate direct PPIs, most of which have been identified *in vitro,* and not within the context of human serum.

#### *The causal protein networks coincide with the co-regulatory network in serum*

We compared the top ranked CPN subnetworks (Table 2, main text), with the previously published serum protein co-regulatory network from the AGES study[2], focusing solely on the network regulators and corresponding target proteins detected by both the 5K and 7K platforms, as the serum co-regulatory network was reconstructed using the 5K aptamer-based platform. We observed a significant overlap between protein clusters in the two network types (Supplementary Table S9 and Supplementary Figure S10A-D). This overlap is evident in two ways: first, many CPNs share the same co-regulatory modules, and second, when a single CPN intersects with multiple co-regulatory modules, these modules frequently belong to the same super-cluster of correlated co-regulatory modules[2]. For example, the PCDH8 CPN subnetwork overlapped with the co-regulatory modules PM16 and PM17 (Supplementary Figure S10A), both in supercluster IV, which is strongly linked to CAD, HF, metabolic syndrome, adiposity, and overall survival[2]. AIPL1 CPN overlapped with PM6, PM7, and PM9 (Supplementary Table S9, Supplementary Figure S10B), all within supercluster II, that is associated with inflammation, CAD, HF, and survival[2]. Additionally, both the C2 and CFB CPNs significantly overlap with PM13 and PM15 from supercluster III (Supplementary Table S9, Supplementary Figure S10C), which has been linked to age-related macular degeneration in the AGES study[3]. Finally, the NUDT21 CPN significantly overlaps with the PM26 and PM27 modules from supercluster V (Supplementary Table S9, Supplementary Figure S10D), which have been previously linked to cardiovascular and metabolic diseases, as well as overall and disease-specific survival[2]. The GABARAP, IZUMO1, FABP3, DCTN2, MRRF, NUDT21, and COL28A1 CPN subnetworks show significant overlaps with the large co-regulatory module PM26 (Supplementary Table S9), which contains 390 proteins, indicating that different CPNs converge into a single, large co-regulatory network. Among the 27 previously identified serum protein co-regulatory subnetworks[2], 12 did not overlap with any of the top ranked CPN subnetworks. Finally, in most instances, when the network regulator aptamer of a specific CPN was available on the 5K platform and assigned to a co-regulatory module, it was part of the overlapping co-regulatory module or super-cluster (Supplementary Table S9), suggesting potential shared genetic influences between these two types of networks. Overall, there is a significant relationship between the circulating CPN and the co-regulatory networks, despite fundamental differences in the methodologies used for their reconstruction.

#### *Additional links between the top-ranked networks and ACVD*

Numerous pathways were enriched among the network regulators from the top ranked subnetworks listed in Table 2 (Supplementary Figure S11). These include pathways previously associated with CVD pathophysiology, such as cellular heat acclimation[4], granulocyte colony-stimulating factor receptor binding[5], farnesylated protein binding[6], vitamin E binding[7] and complement system-related functions[8], among others. Functional enrichment analysis of the CPN target members within the top-ranked networks identified numerous pathways that were overrepresented across various subnetworks (Supplementary Table S10, Supplementary Figure S12). While these pathways differ from those identified for the combined group of network regulators, both sets share enrichment in cellular response to heat shock. Finally, we explored known functional and physical protein-protein interactions among the top ranked serum protein network regulators using the STRING database[9] and observed a significant enrichment in interaction levels (P = 0.0473), indicating that these network regulators interact more frequently than expected by chance. This analysis reflects some of the previously identified interactions shown in Figure 6A and offers additional insights, such as interactions involving the top ranked network regulator ITIH3, which shows physical connections with KLKB1, APOA5, and AFM (Supplementary Figure S13).

Several top-ranked network regulators have previously been linked to myocardial infarction (MI) and related traits, including the proinflammatory protein ITIH3, which emerged as the leading subnetwork (Table 2). The ITIH3 subnetwork comprises 11 protein members and is enriched in the ubiquitin-mediated proteolysis pathway (Supplementary Table S10). Both the network regulator and the corresponding eigenprotein were associated with incident MI and related traits, except for T2D (Table 2, Figure 5). Interestingly, the data-driven reconstruction of the CPN reveals that the roots of the global network regulating a diverse array of network targets converge on ITIH3 at the base of the network (Figure 3), suggesting its involvement in many regulatory pathways. Notably, a functional genetic variant in the *ITIH3* gene has previously been linked to MI and the protein is expressed in vascular smooth muscle cells and macrophages within human atherosclerotic lesions[10].

Other network regulators from the top ranking subnetworks previously linked to risk of ACVD include: 1) Genetic variant across the *KEAP1* gene, show strong genetic links to low-density lipoprotein (LDL) cholesterol levels[11], as well as familial hypercholesterolemia and ischemic heart disease in the FinnGen study[12]. 2) Genetic risk variants in the *C2* gene are associated with CAD[13]. 3) The gene encoding GABARAP, is linked to genetic risk of CVD (excluding rheumatic disease)[12]. 4) Genetic variants near the *KLKB1* gene, are linked to venous thromboembolism[12]. 5) Variants within the gene encoding *APOA5*, are associated with coronary atherosclerosis and coronary revascularization[12], as well as hyperlipoproteinemia in multiple studies[14]. Additionally, APOA5 demonstrated a robust causal relationship with both MI and MetS in the AGES study, supported by results from both colocalization and MR analyses (Supplementary Table S12). 6) The gene encoding *PTPN11*, is linked to major coronary heart disease events[12], and right ventricular end-diastolic volume in HF[15]. 7) The gene encoding *CFB*, is linked to genetic risk variants of CAD[16], while 8) the colocalized genes on chromosome 11 encoding C11orf49 or LRP4, are associated with cardiomyopathy[12] or venous thromboembolism[17], respectively. Interestingly, both C11orf49 and LRP4 were identified as having a causal relationship with MI in this study, based on findings from both the MR and colocalization analyses (Supplementary Table S12). However, since their *cis* regions overlap, from which the genetic instruments are derived, it is not possible to determine whether one or both proteins are causal effectors. HSPA1A and HSPA1B have been linked to T2D and its microvascular complications through both MR and colocalization analyses[18]. In our study, HSPA1A was found to be causally related to MetS, as indicated by both MR and colocalization analyses (Supplementary Table S12). Several of the top-ranked network regulators, including for instance DDX39B, have not been previously linked directly to MI and/or related traits. In our MR and colocalization analyses, DDX39B was found to be causally associated with T2D (Supplementary Table S12). The regulator protein DDX39B has been shown to play a role in modulating the NF-κB response[19], a key pathway involved in immune and inflammatory responses. This modulation may have implications for diseases like atherosclerosis and diabetes, where NF-κB signaling is frequently dysregulated causing inflammation in the vascular wall, insulin resistance and/or beta-cell dysfunction[20].

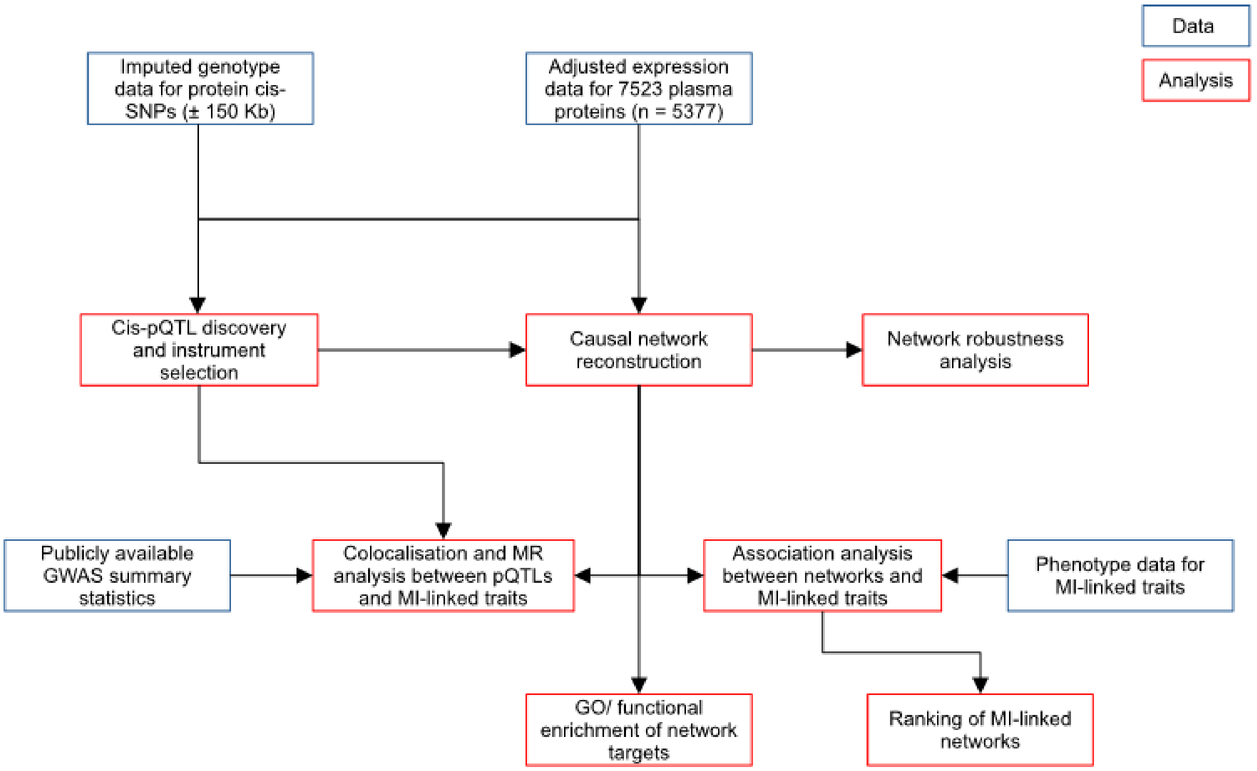


**Supplementary Figure S1**. A flowchart illustrating the reconstruction and analysis of the circulating causal protein networks (CPNs) in relation to incident myocardial infarction (MI) and associated phenotypes. An alternate version of the study overview is presented in Figure 1 of the main text.

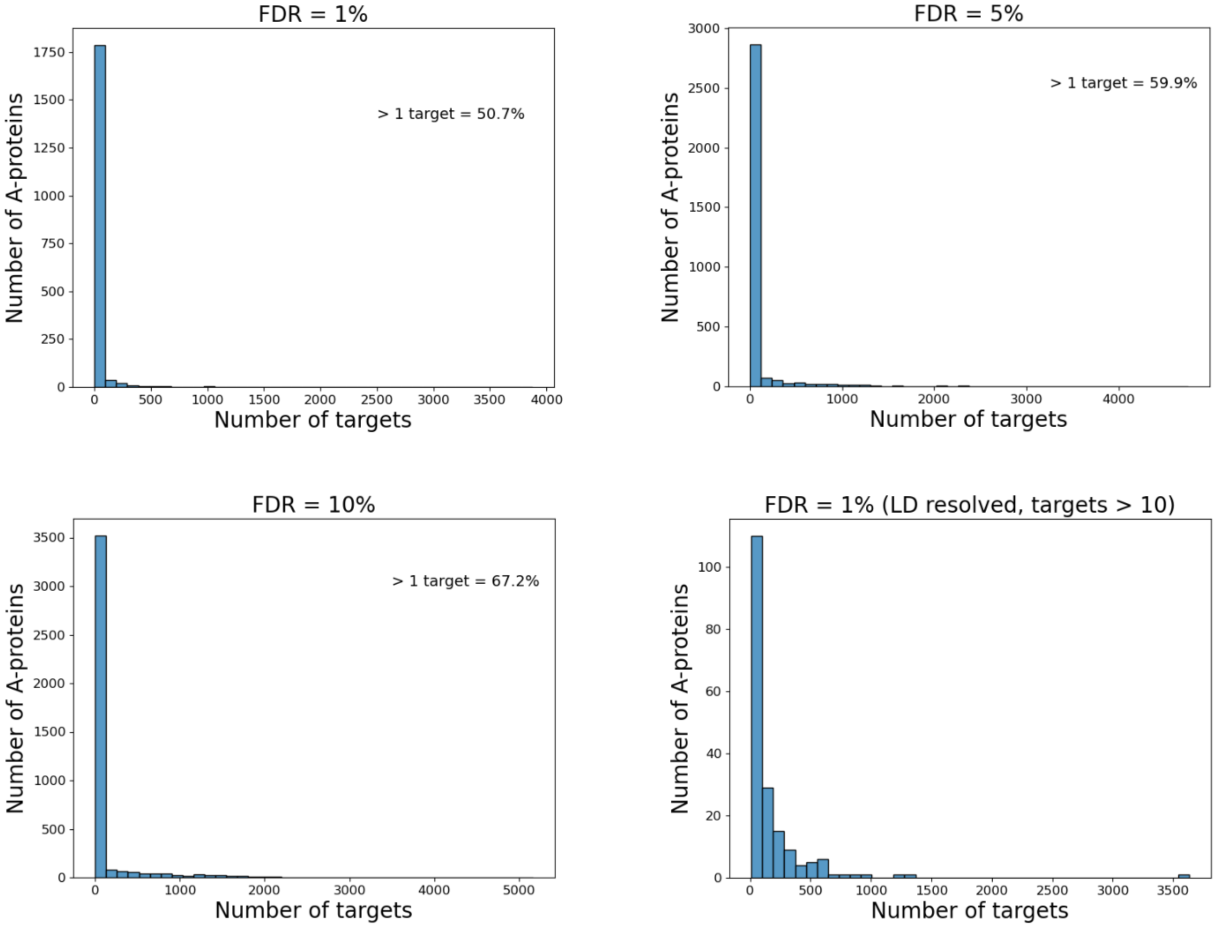


**Supplementary Figure S2**. Histograms of network target distributions are shown at various FDR thresholds, followed by the target distribution in the CPN based on a minimum of 10 unique protein members per subnetwork.

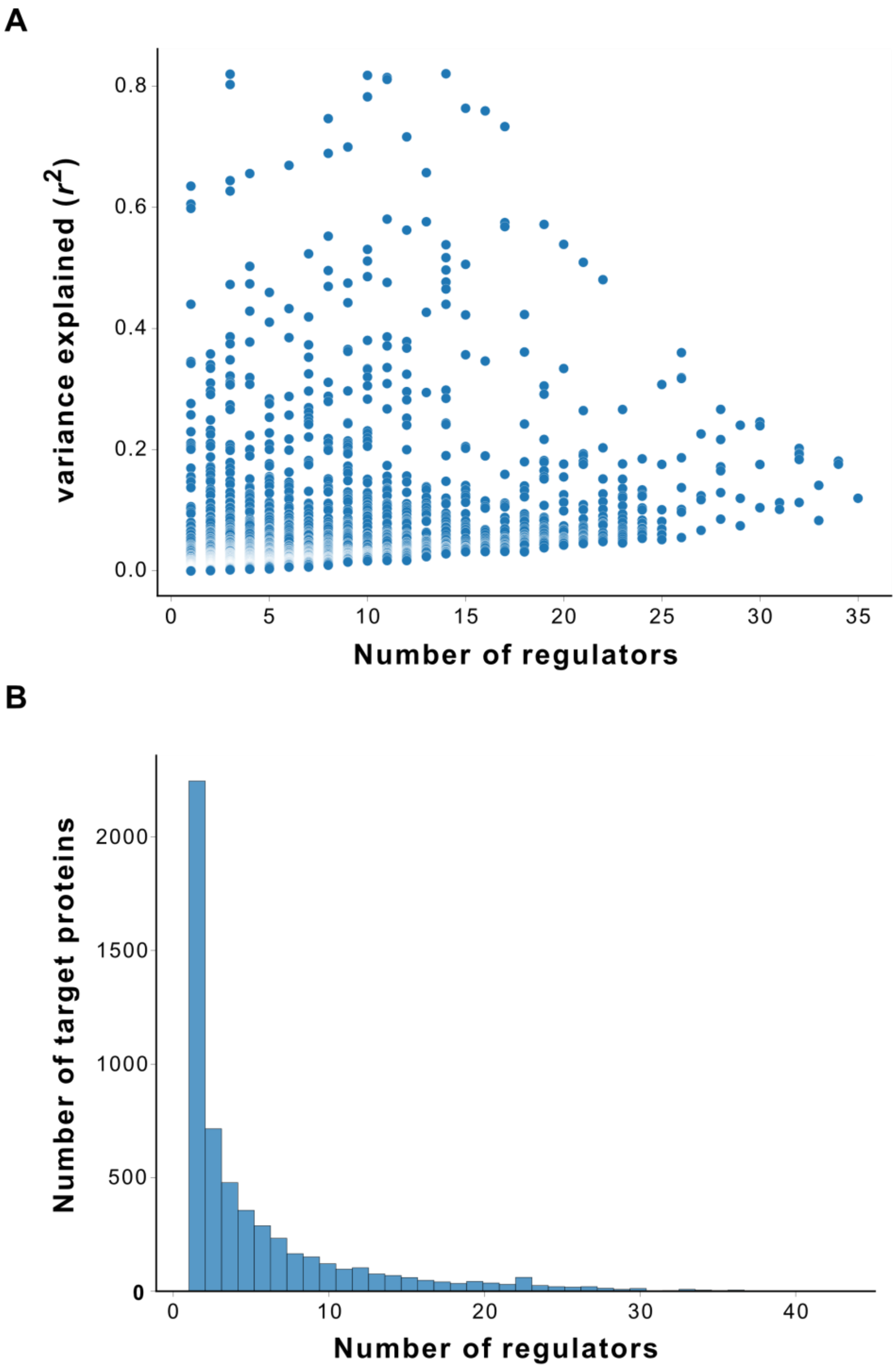


**Supplementary Figure S3**. **A**. The proportion of variance in protein expression (adjusted $r^2$) explained by *cis*-acting pQTLs for the regulatory proteins' impact on 5,459 target proteins in the CPN. **B.** The number of network regulators each of the 5,499 target proteins has.

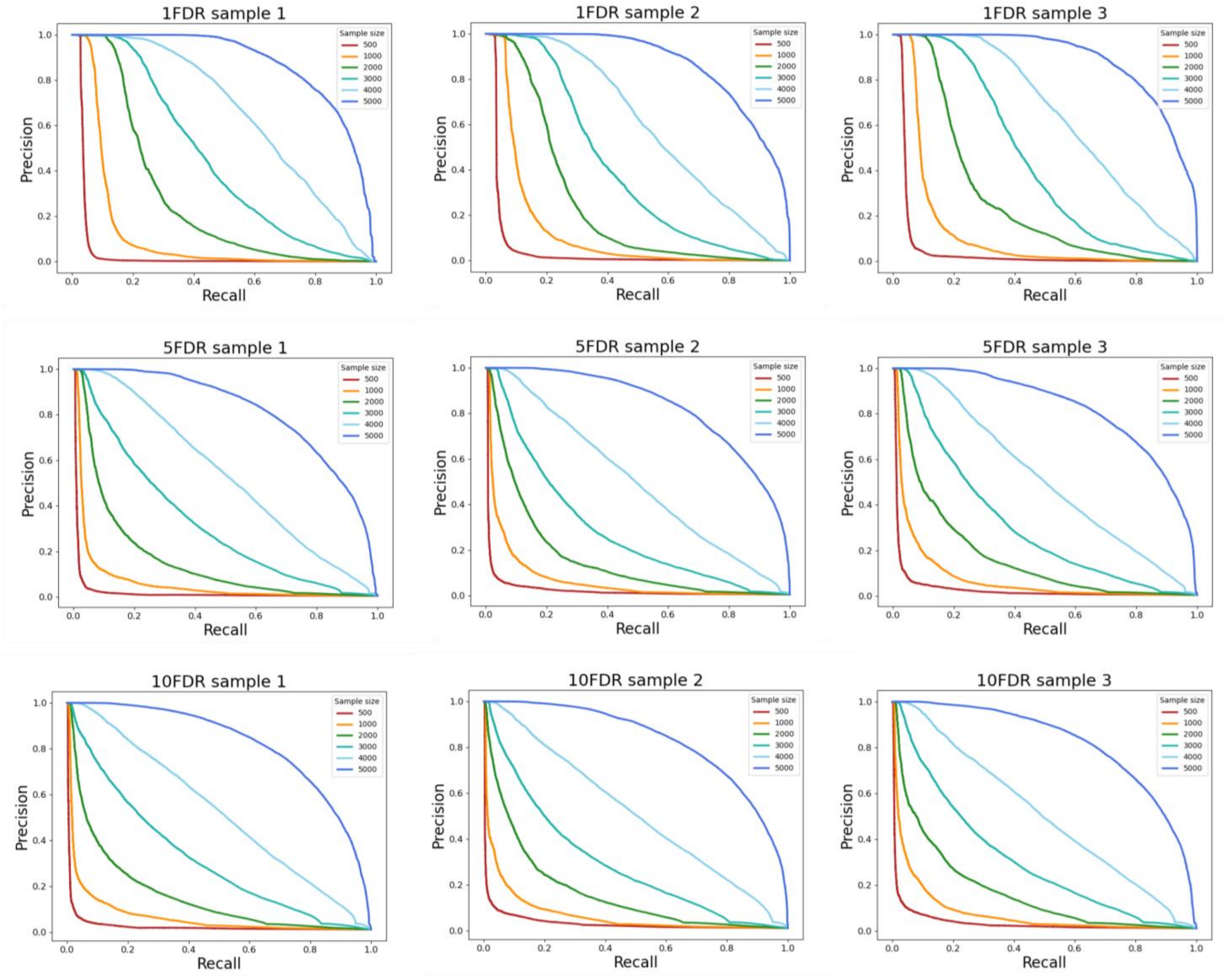


**Supplementary Figure S4**. Precision-Recall curves were generated for sub-sampled networks and compared to networks using the full set of samples. Three random sub-samples of AGES participants were selected at different thresholds: 500, 1000, 2000, 3000, 4000, and 5000 samples, and networks reconstructed for each of these sub-sample test sets. The network generated from the full sample set served as the ground truth, filtered at 5%, and 10% FDR. The network filtered at 1% FDR is presented as Figure 3B in the main text. These ground truth networks were represented as flattened matrices (i,j), where i = network regulator and j = target, with a value of 1 indicating an edge and 0 indicating no edge. Receiver operating characteristic (ROC) AUC and precision-recall were calculated for each sub-sampled network and compared against the ground truth networks.

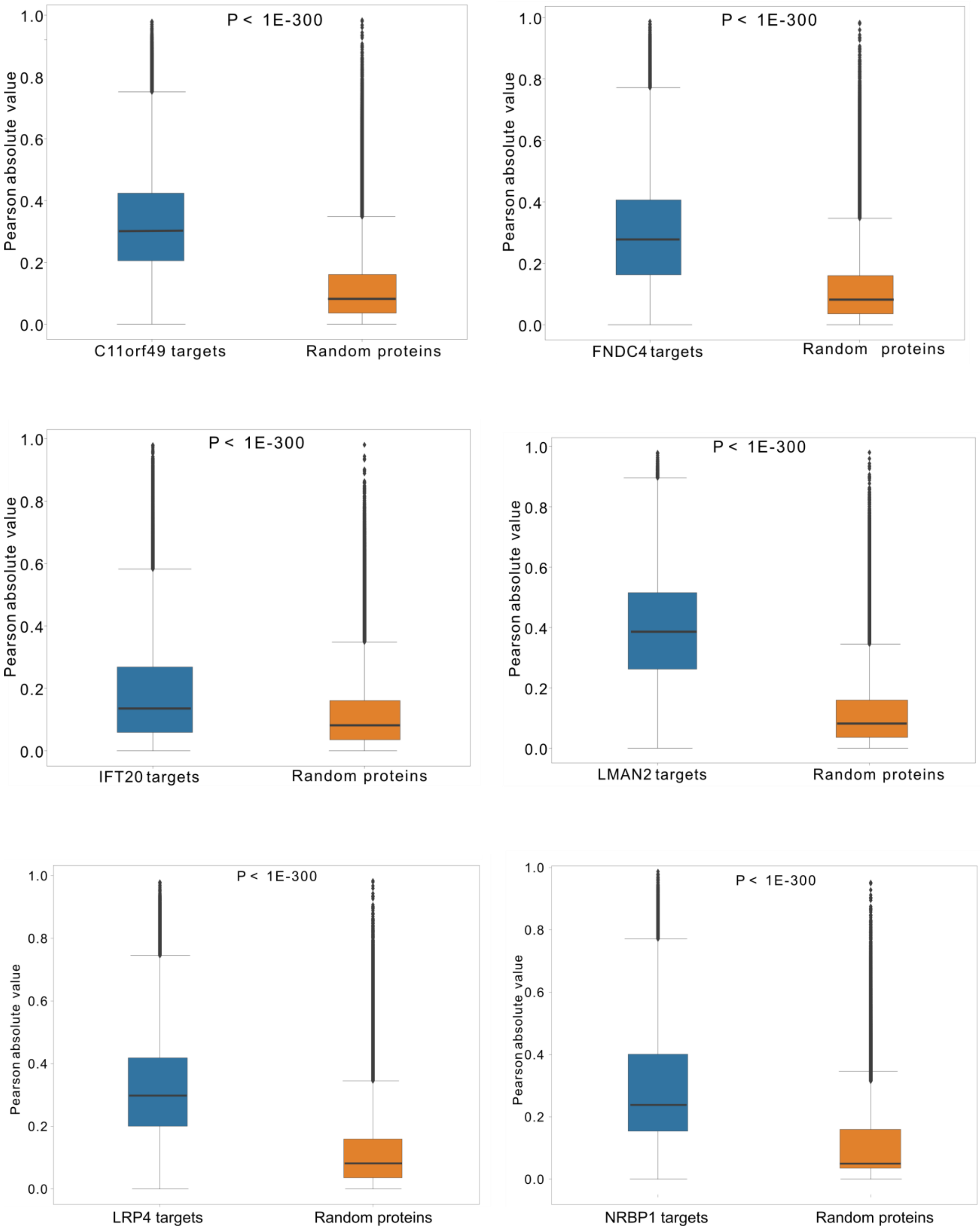

P < 1E-300
Pearson absolute value
1.0
0.8
0.6
0.4
0.2
0.0
C11orf49 targets
Random proteins
P < 1E-300
Pearson absolute value
FNDC4 targets
Random proteins
P < 1E-300
Pearson absolute value
IFT20 targets
Random proteins
P < 1E-300
Pearson absolute value
LMAN2 targets
Random proteins
P < 1E-300
Pearson absolute value
LRP4 targets
Random proteins
P < 1E-300
Pearson absolute value
NRBP1 targets
Random proteins

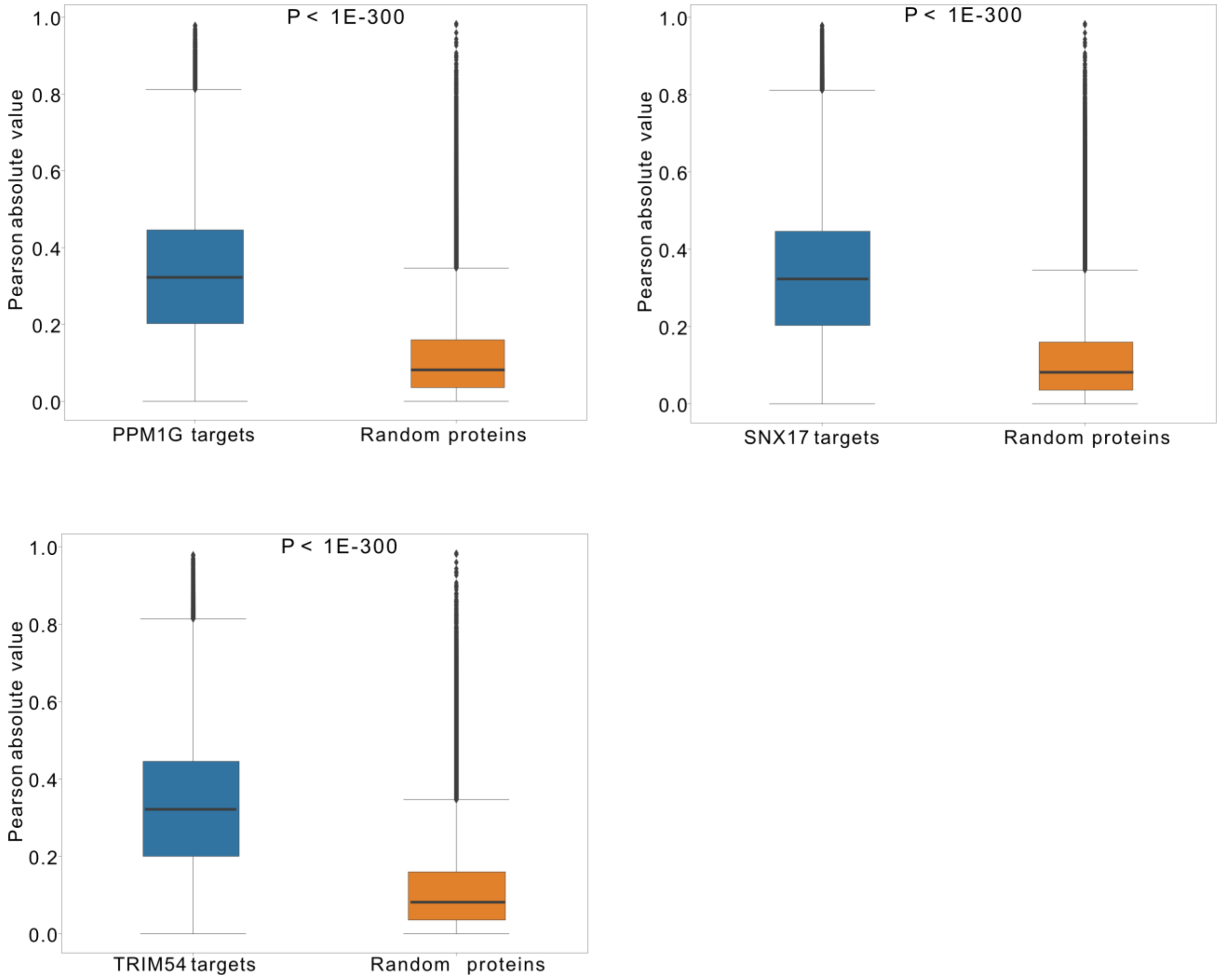


**Supplementary Figure S5**. Box plots of a pairwise Pearson correlations between all proteins in a given CPN (blue boxes) compared to pairwise correlations between random proteins of the same size (yellow brown boxes). The P-value is a Kruskal Wallis test for the predicted vs random distributions.

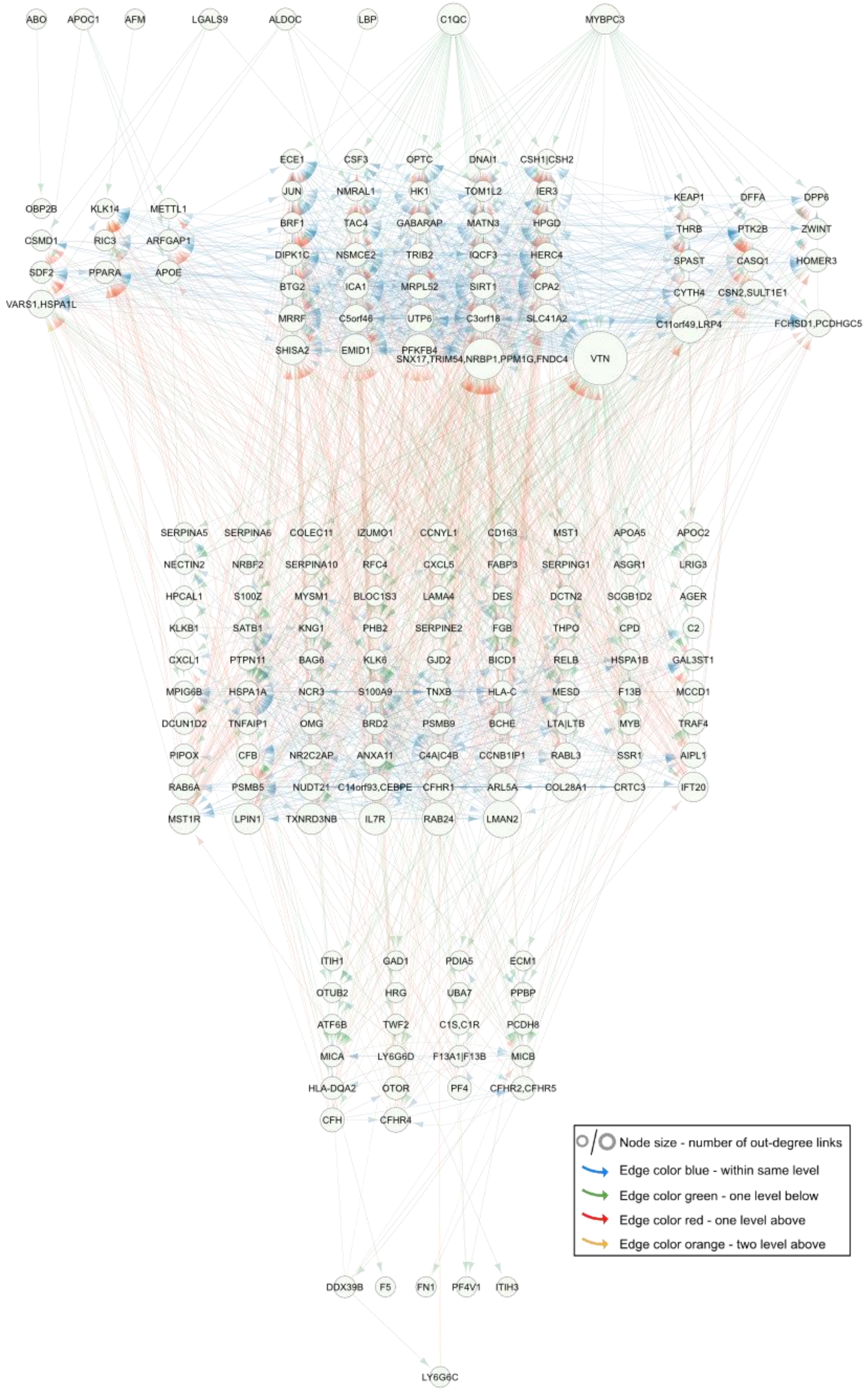


**Supplementary Figure S6**. Network visualization of causal interactions among the 185 CPNs with more than 10 targets (FDR = 1%), with no edges removed. Refer to the comparison with the DAG Bayesian network in Figure 3 of the main text.

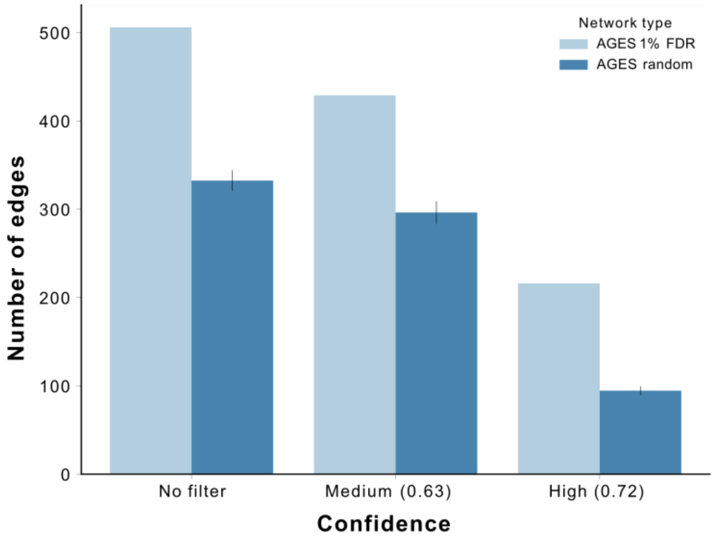


**Supplementary Figure S7**. The CPN networks identified in the AGES study were compared against 289,112 protein-protein interactions (PPIs) sourced from the human integrated protein-protein interactions reference database at varying confidence thresholds (see Methods).

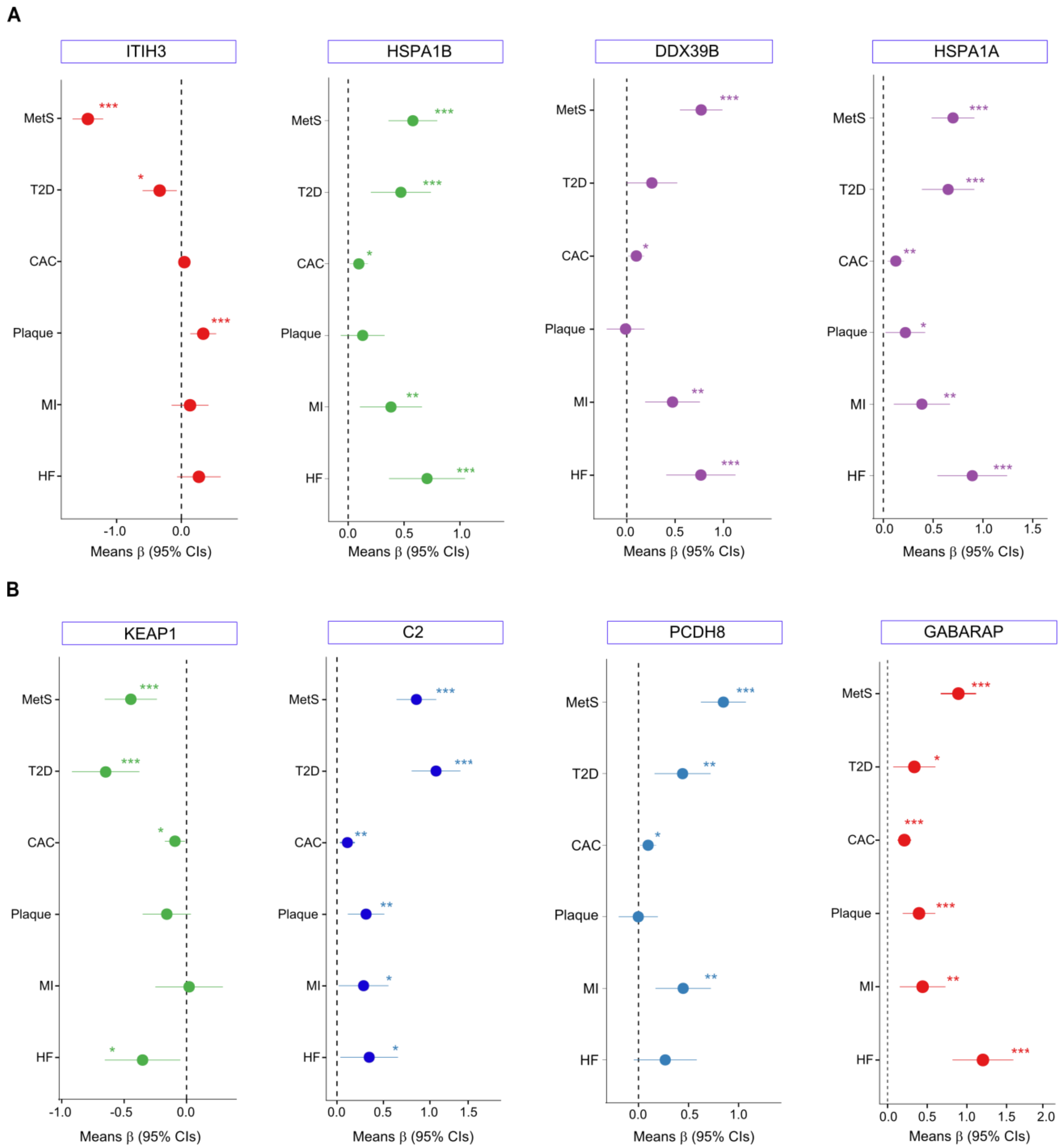
A
ITIH3
HSPA1B
DDX39B
HSPA1A
MetS
T2D
CAC
Plaque
MI
HF
Means β (95% CIs)
B
KEAP1
C2
PCDH8
GABARAP

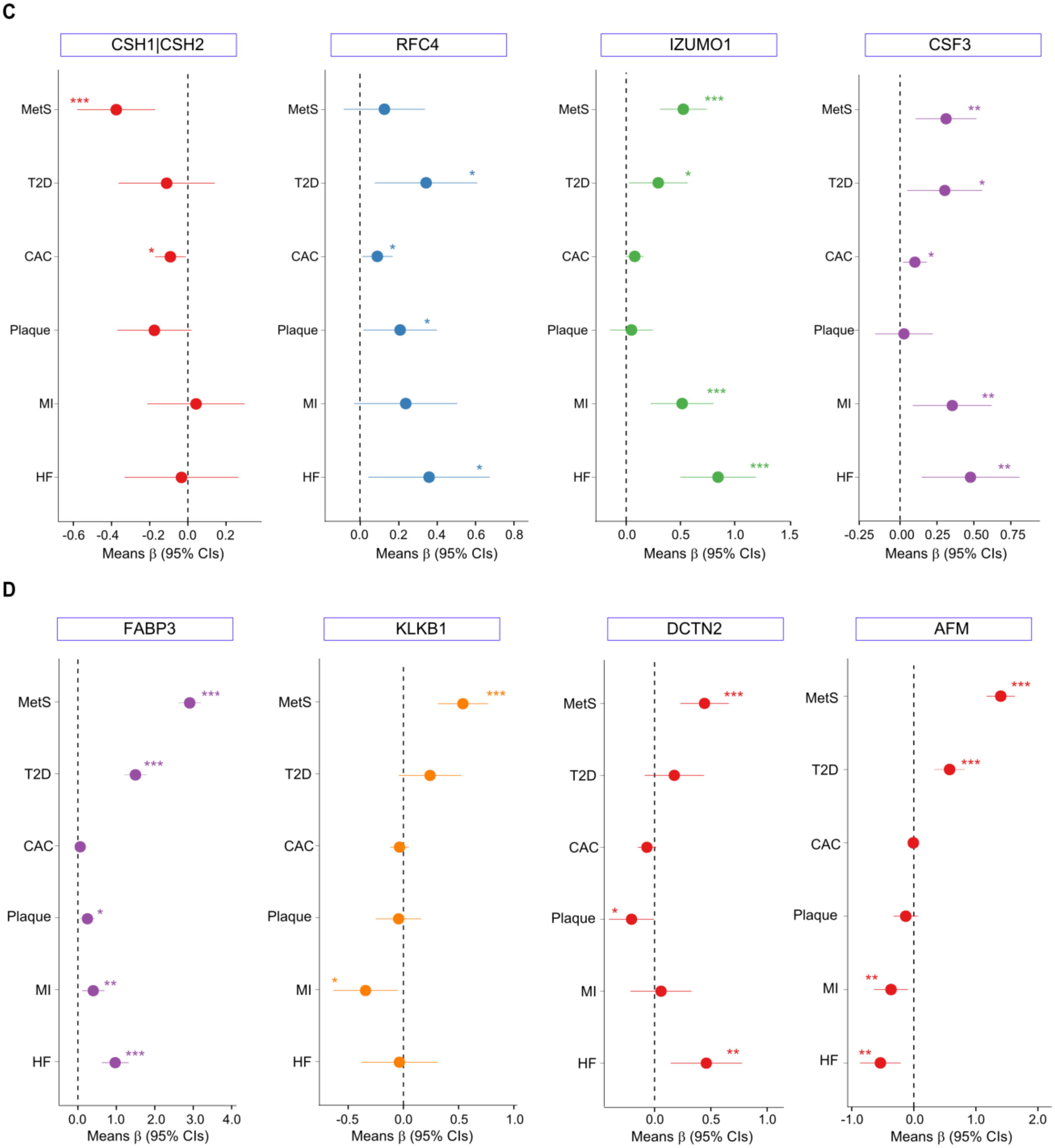
C
CSH1|CSH2
RFC4
IZUMO1
CSF3
MetS
T2D
CAC
Plaque
MI
HF
-0.6 -0.4 -0.2 0.0 0.2
0.0 0.2 0.4 0.6 0.8
0.0 0.5 1.0 1.5
-0.25 0.00 0.25 0.50 0.75
Means β (95% CIs)
D
FABP3
KLKB1
DCTN2
AFM
0.0 1.0 2.0 3.0 4.0
-0.5 0.0 0.5 1.0
0.0 0.5 1.0
-1.0 0.0 1.0 2.0

E

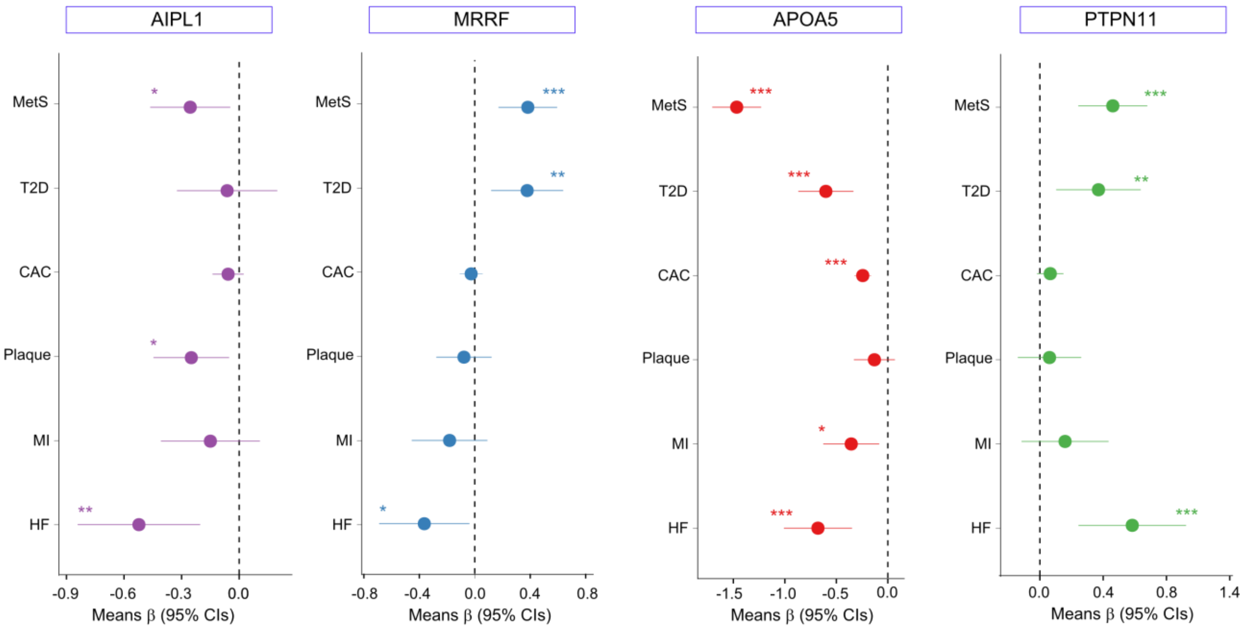
AIPL1
MRRF
APOA5
PTPN11
MetS
T2D
CAC
Plaque
MI
HF
*
**
***
-0.9
-0.6
-0.3
0.0
-0.8
-0.4
0.0
0.4
0.8
-1.5
-1.0
-0.5
0.0
0.0
0.4
0.8
1.4
Means β (95% CIs)

F

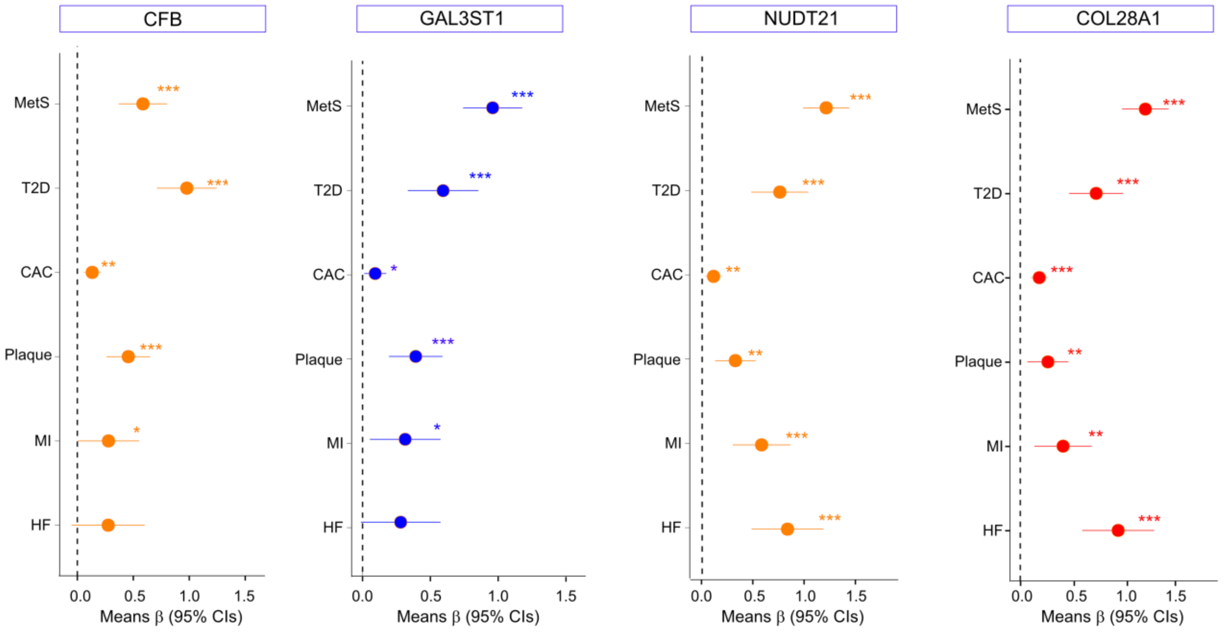
CFB
GAL3ST1
NUDT21
COL28A1
MetS
T2D
CAC
Plaque
MI
HF
*
**
***
0.0
0.5
1.0
1.5
Means β (95% CIs)

G

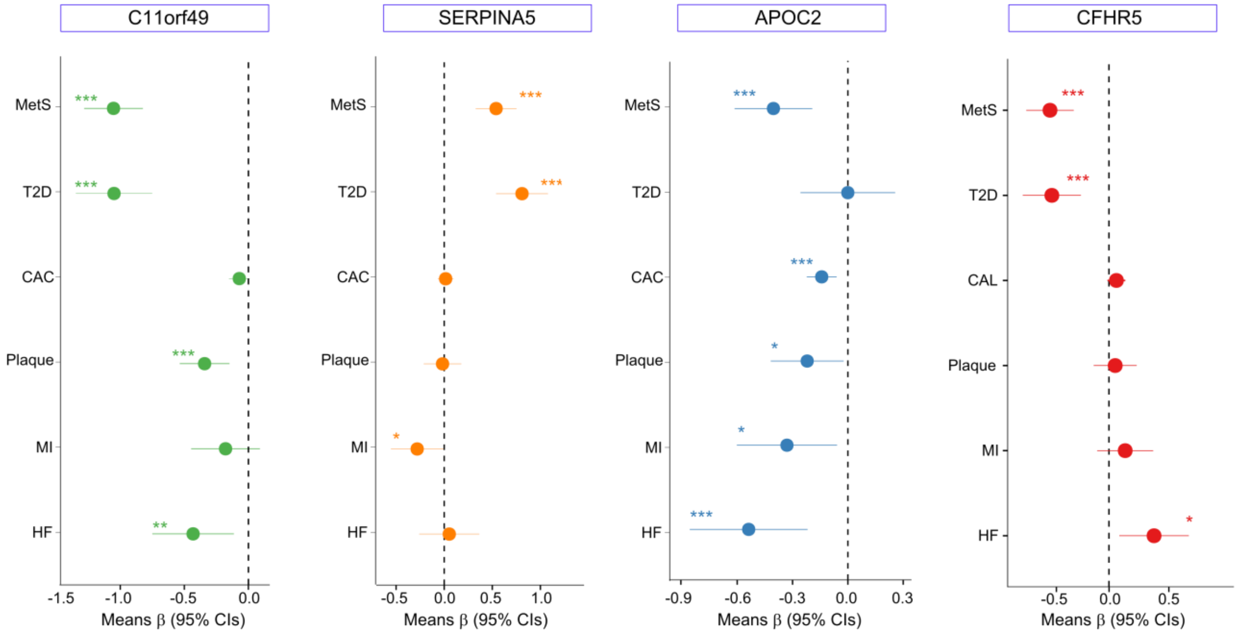
C11orf49
SERPINA5
APOC2
CFHR5
MetS
T2D
CAC
Plaque
MI
HF
CAL
-1.5
-1.0
-0.5
0.0
0.5
1.0
-0.9
-0.6
-0.3
0.3
Means β (95% CIs)

H

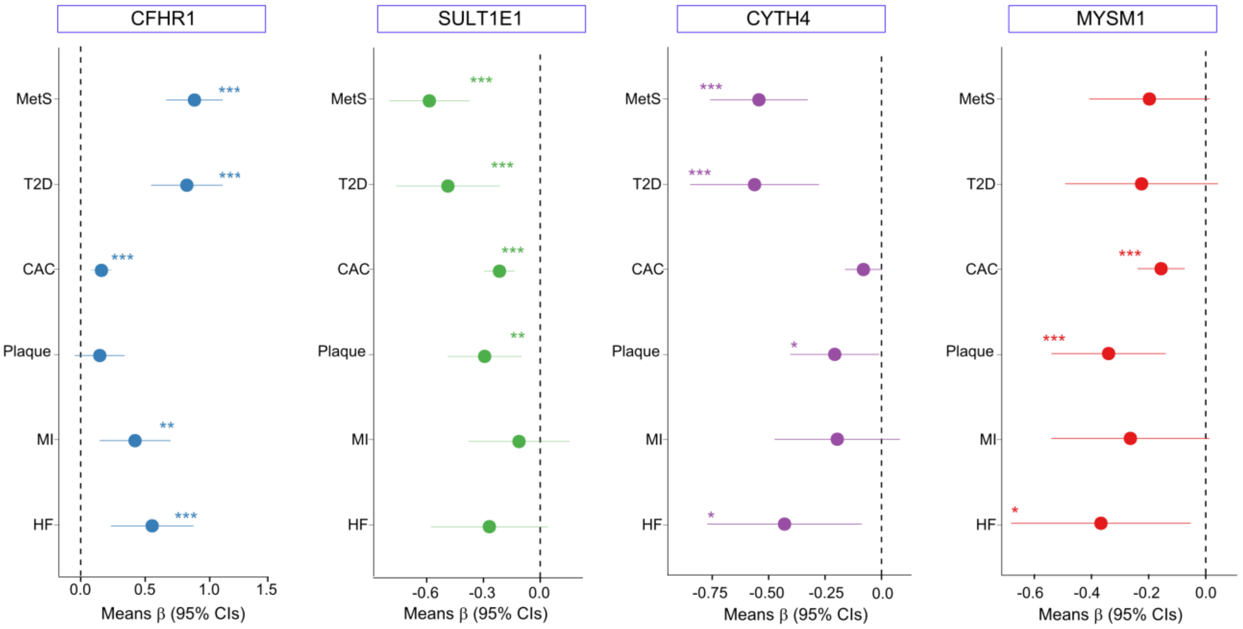
CFHR1
SULT1E1
CYTH4
MYSM1
MetS
T2D
CAC
Plaque
MI
HF
0.0
0.5
1.0
1.5
-0.6
-0.3
-0.75
-0.50
-0.25
-0.4
-0.2
Means β (95% CIs)

**I**

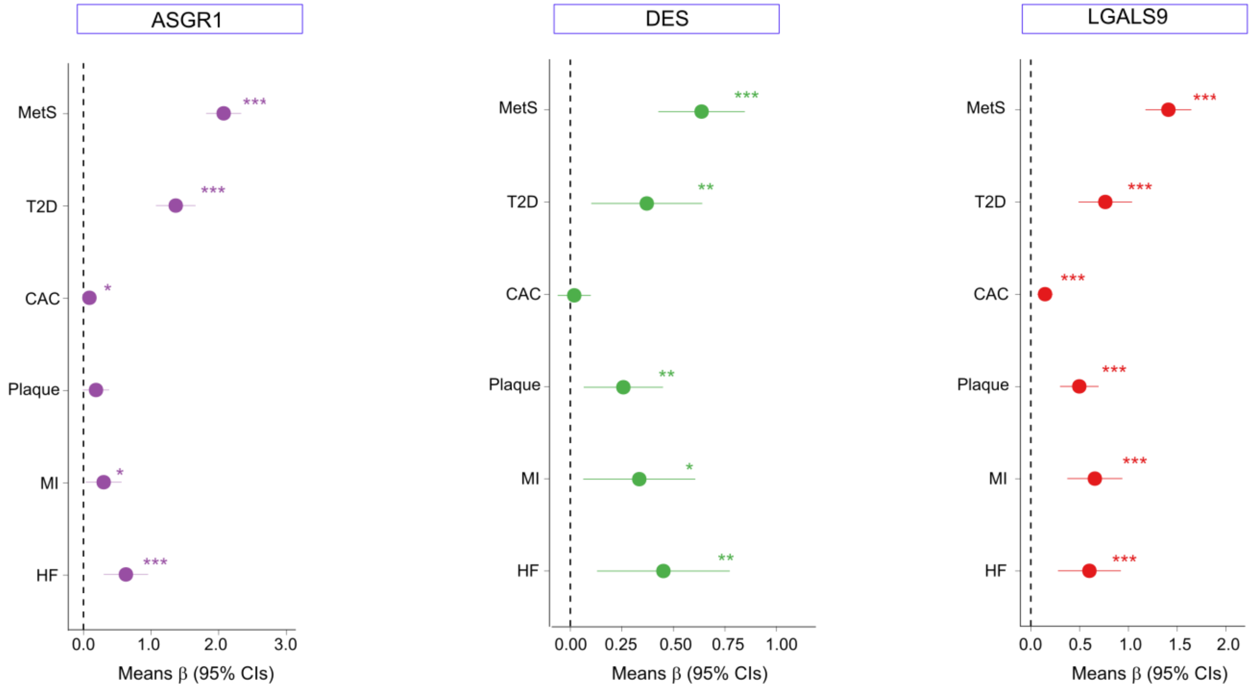


**Supplementary Figure S8.** The difference in various outcome values between the fifth (top) and first (bottom) quintiles of the network regulators (**A-I**) from the top ranking CPNs. For each aptamer, we split the AGES cohort into quintile groups and calculate associations between the group (treated as a factor variable) and an assortment of outcomes. In the case of a continuous outcome, we use ordinary regression, and in the case of binary outcomes we use logistic regression for prevalent disease and Cox proportional-hazards model fitted to the box cox transformed serum proteomics data. All statistical results are obtained using linear models in the case of continuous outcomes and generalized linear models for binary outcomes. The models were fit using the outcomes as dependent variables and protein quintiles as predictor variables along with any adjustment variables including age or sex. The protein quintiles are treated as factor variables so there are no underlying assumptions regarding linear effects or other functional forms. Continuous outcomes are standardized prior to model fitting so coefficient estimates should be interpreted on the standard deviation scale, i.e. an estimated mean difference of 1 between protein quintiles translates to a one-standard-deviation difference between groups after adjusting for other included variables. The expected means are obtained as linear predictions from the fitted models along with the fitted confidence intervals around the mean. The linear predictions for qualitative phenotypes are shown on the log-odds scale. The difference between the 5th and 1st protein quintiles (or any other quintiles) is obtained as the expected marginal difference between those groups after adjusting for any other included variables with unadjusted p-values. As such, for continuous outcomes they are the optimal linear estimator with corresponding confidence intervals and p-values, but for discrete outcomes they are obtained using commonly applied asymptotic approximations. MetS, metabolic syndrome; T2D, type two diabetes; CAC, coronary artery calcium; Plaque, carotid plaque severity score; MI, incident myocardial infarction; HF, incident heart failure. ***(P-value < 0.001, two-sided), **(P-value < 0.01, two-sided), *(P-value < 0.05, two-sided).

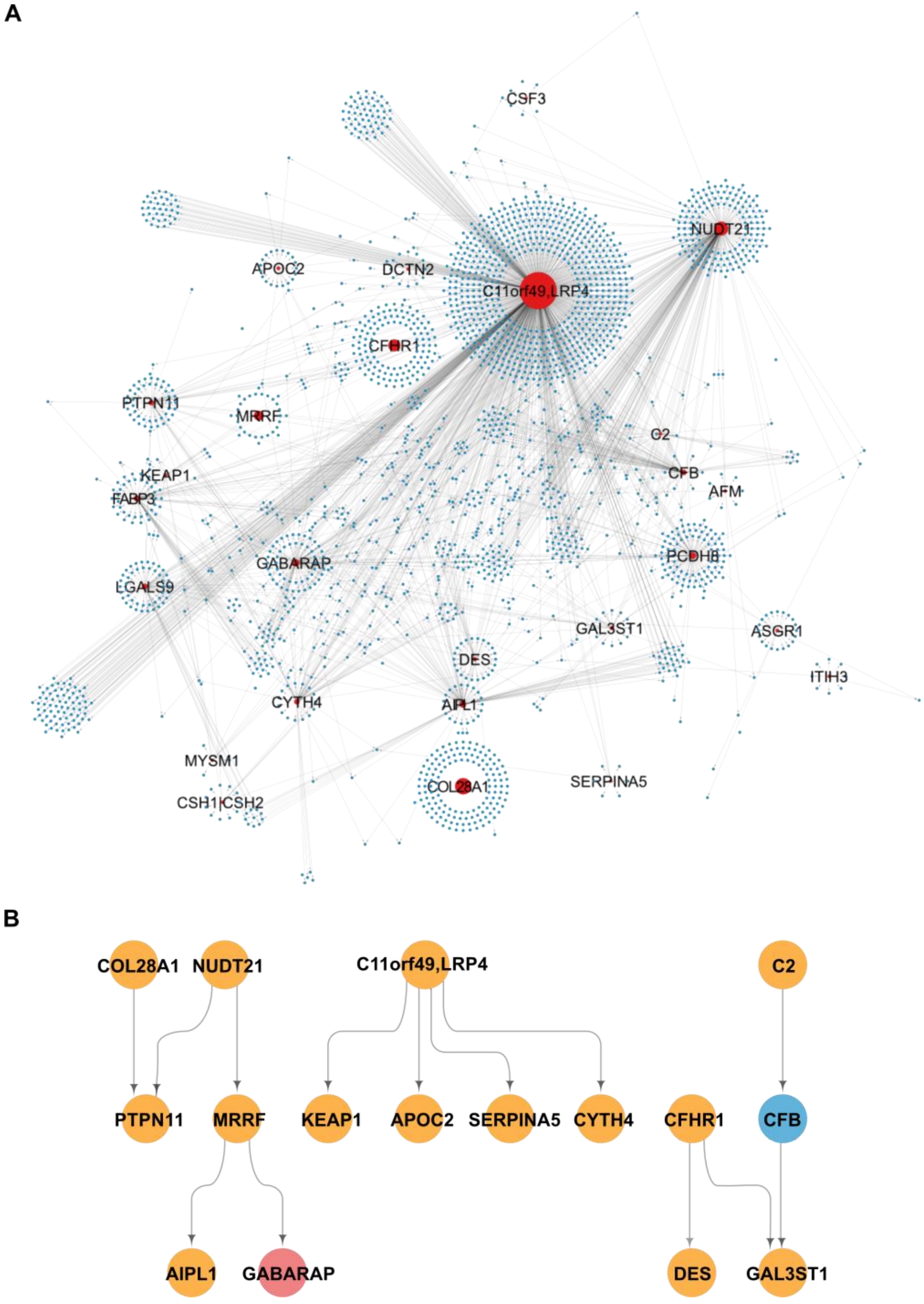


**Supplementary Figure S9**. **A**. Network visualization of top ranked CPNs linked to MI-linked traits (arbitrary ranking score > 6) where network eigen-protein variance explained > 30%. Red nodes represent regulatory A-proteins and blue nodes represent target B-proteins. **B**. A hierarchical representation of regulatory A-proteins, where color indicates the degree of association with incident MI. Blue signifies no association, yellow indicates that either the eigen-protein or A-protein is associated, and red denotes that both the eigen-protein and A-protein are associated. The eigen-protein PC1 was required to explain at least 30% of the variance (refer to the top-ranked networks for incident MI and HF related traits in Supplementary Tables S7 and S8).

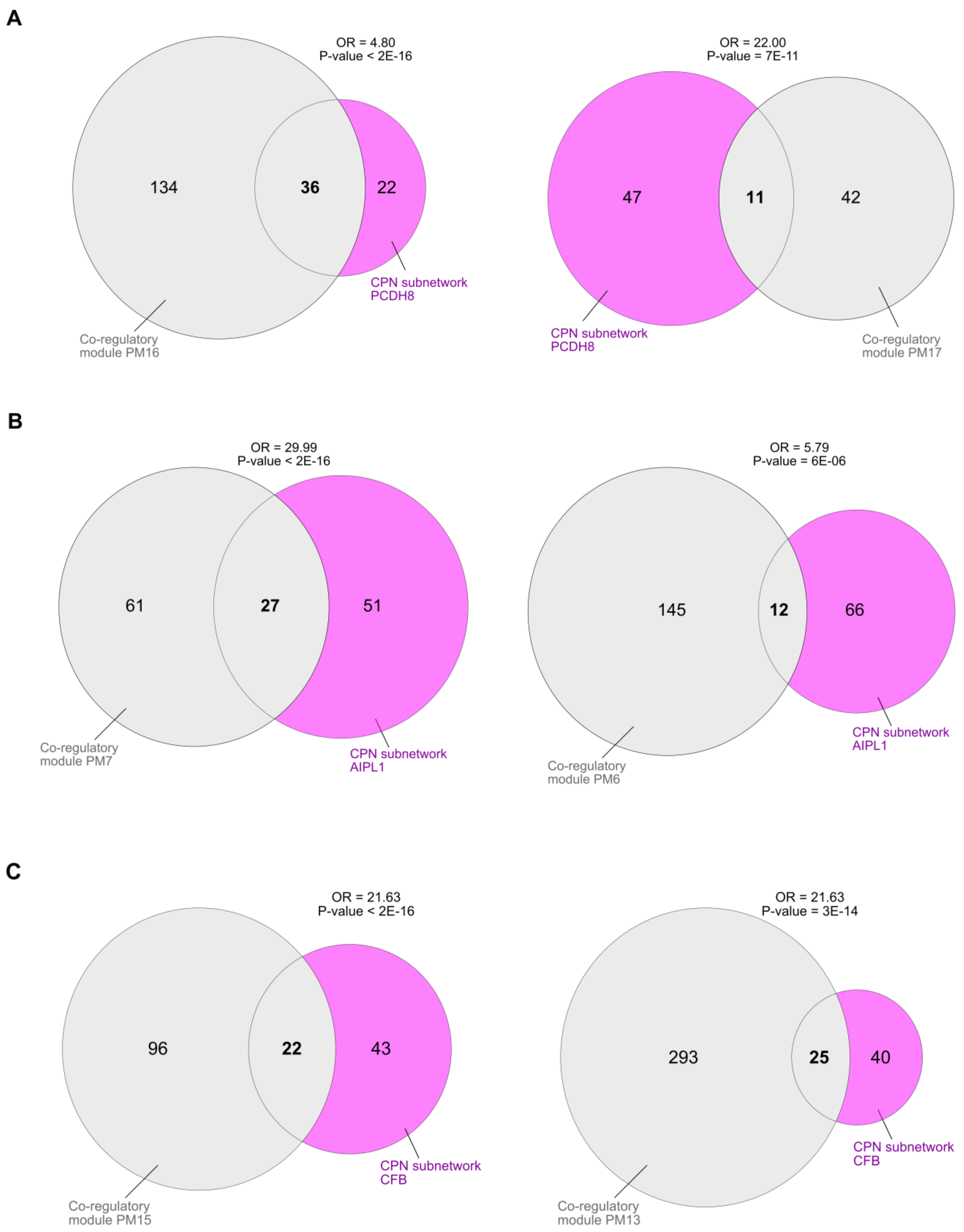
A
OR = 4.80
P-value < 2E-16
134
36
22
CPN subnetwork PCDH8
Co-regulatory module PM16
OR = 22.00
P-value = 7E-11
47
11
42
CPN subnetwork PCDH8
Co-regulatory module PM17
B
OR = 29.99
P-value < 2E-16
61
27
51
Co-regulatory module PM7
CPN subnetwork AIPL1
OR = 5.79
P-value = 6E-06
145
12
66
CPN subnetwork AIPL1
Co-regulatory module PM6
C
OR = 21.63
P-value < 2E-16
96
22
43
CPN subnetwork CFB
Co-regulatory module PM15
OR = 21.63
P-value = 3E-14
293
25
40
CPN subnetwork CFB
Co-regulatory module PM13

**D**

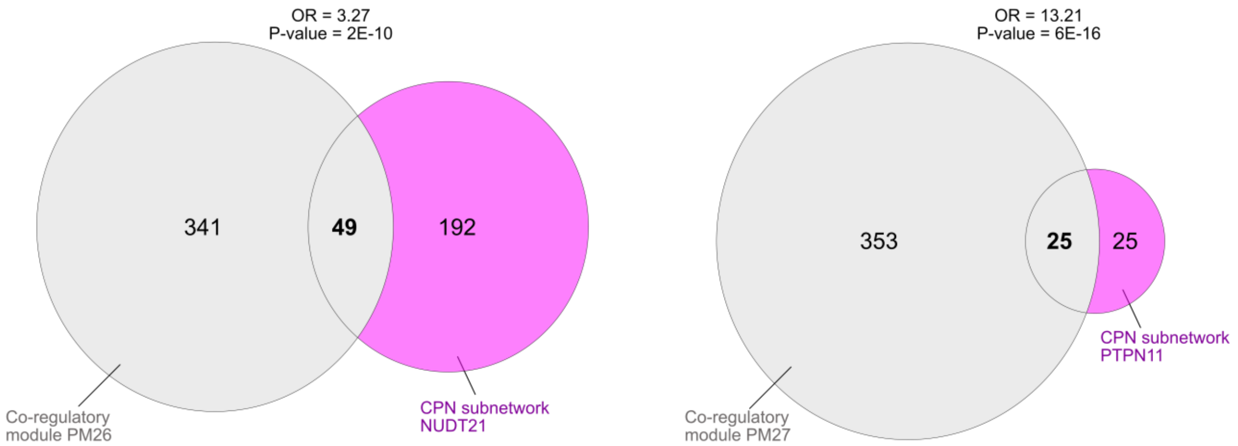


**Supplementary Figure S10**. Examples (**A**-**D**) of Fisher's exact test results for the overlap between top-ranked circulating causal protein networks (CPN) in Table 2 of the main text, and the serum protein co-regulatory networks (PM) from Emilsson et al.[2]. For this comparison we focused solely on the proteins detected by both the 5K and 7K aptamer platforms. Complete details of these overlaps can be found in Supplementary Table S9.

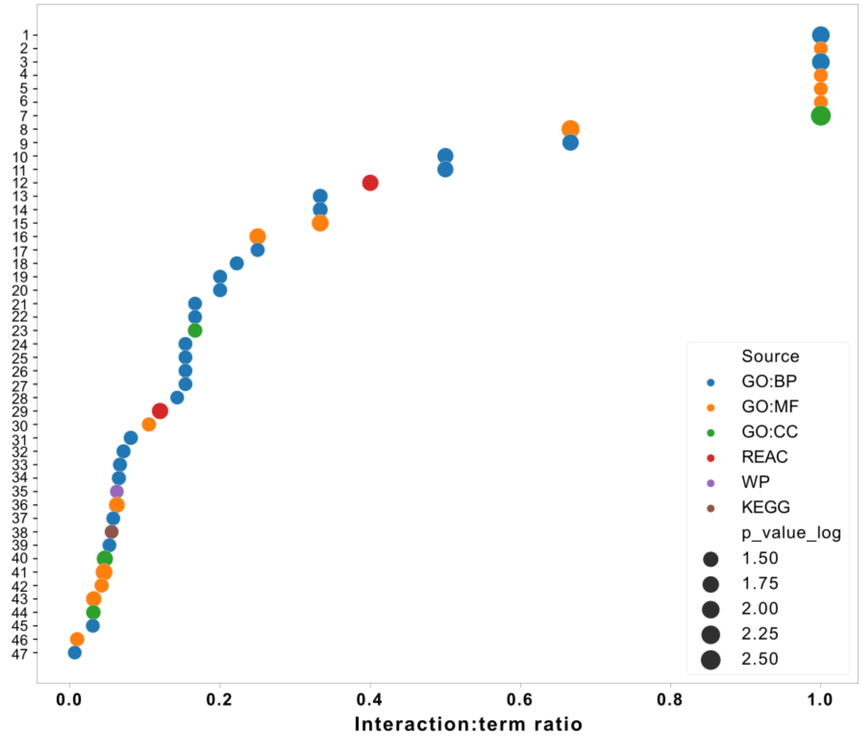


1. Cellular heat acclimation
2. Granulocyte colony-stimulating factor receptor binding
3. Heat acclimation
4. Farnesylated protein binding
5. Denatured protein binding
6. Vitamin E binding
7. Classical-complement-pathway C3/C5 convertase complex
8. C3HC4-type RING finger domain binding
9. Positive regulation of microtubule nucleation
10. Positive regulation of nucleotide-binding oligomerization domain containing 2 signaling pathway
11. Positive regulation of nucleotide-binding domain, leucine rich repeat containing receptor signaling pathway
12. Activation of C3 and C5
13. Regulation of nucleotide-binding oligomerization domain containing 2 signaling pathway
14. Regulation of microtubule nucleation
15. ATP-dependent protein disaggregase activity
16. Protein-containing complex destabilizing activity
17. Regulation of nucleotide-binding domain, leucine rich repeat containing receptor signaling pathway
18. Positive regulation of tumor necrosis factor-mediated signaling pathway
19. Negative regulation of inclusion body assembly
20. Nucleotide-binding oligomerization domain containing 2 signaling pathway
21. Regulation of mitotic spindle assembly
22. Regulation of spindle assembly
23. Aggresome
24. Regulation of Rac protein signal transduction
25. Microtubule nucleation
26. Nucleotide-binding domain, leucine rich repeat containing receptor signaling pathway
27. Positive regulation of microtubule polymerization
28. Regulation of inclusion body assembly
29. HSP90 chaperone cycle for steroid hormone receptors (SHR) in the presence of ligand
30. ATP-dependent protein folding chaperone
31. Positive regulation of proteasomal ubiquitin-dependent protein catabolic process
32. Positive regulation of protein polymerization
33. Positive regulation of ubiquitin-dependent protein catabolic process
34. Positive regulation of proteasomal protein catabolic process
35. Complement and coagulation cascades
36. Histone deacetylase binding
37. Mitotic spindle organization
38. Spliceosome
39. Positive regulation of proteolysis involved in protein catabolic process
40. Blood microparticle
41. ATP hydrolysis activity
42. Unfolded protein binding
43. ATP-dependent activity
44. Nuclear speck
45. Glycerolipid metabolic process
46. Enzyme binding
47. Cellular component organization

**Supplementary Figure S11**. A functional enrichment analysis conducted on the 25 network regulators from the top ranking CPNs. The bubble plot displays the ratio of interaction size to term size, with the dot size representing the -log10 P-value. Significant enrichment was defined as FDR < 5%.

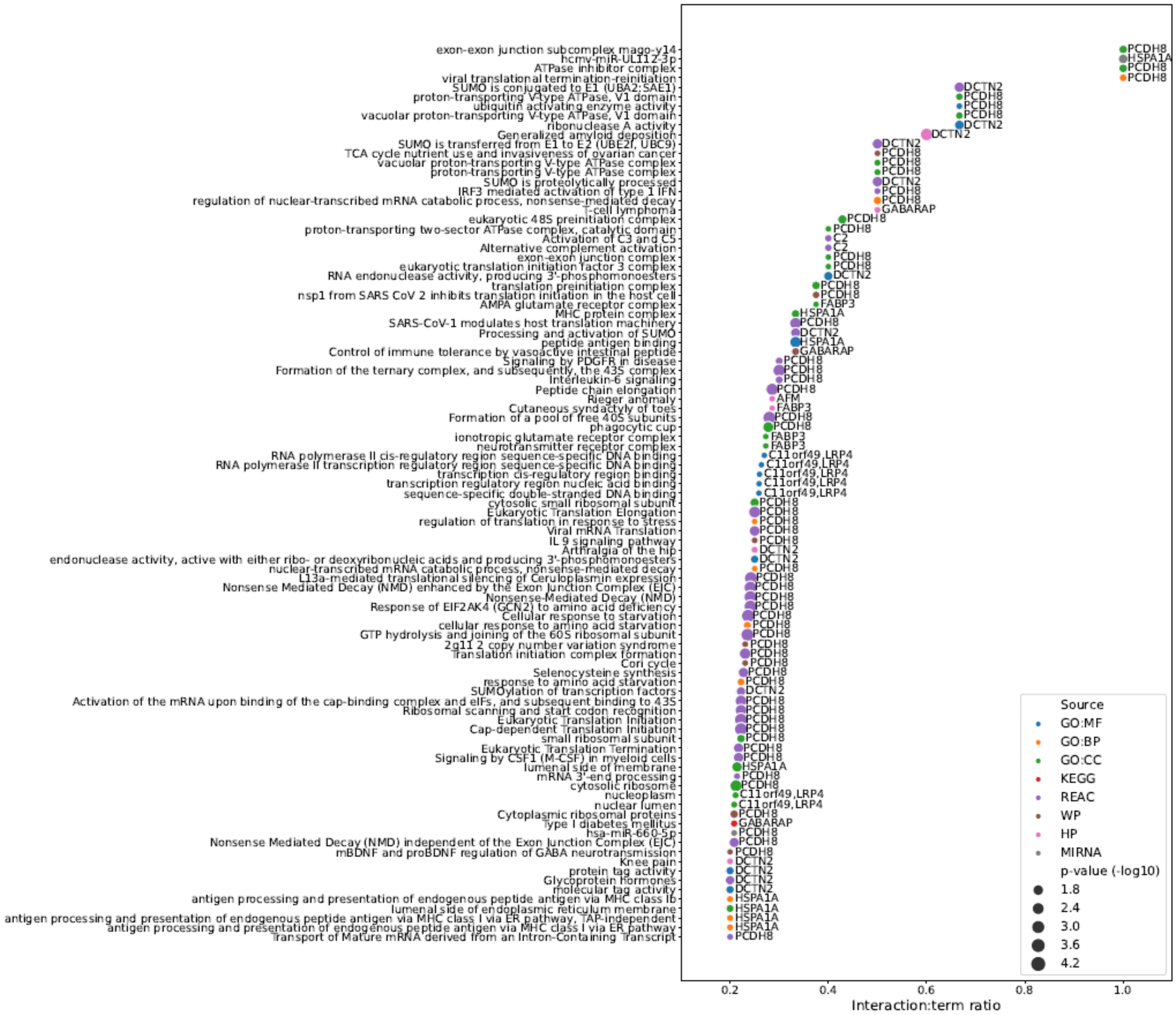


**Supplementary Table S12**. GO enrichments for all targets within each top-ranked CPN subnetwork were analyzed using g:Profiler, similar to that in Figure S11, with the respective networks labeled on the plot. These enrichments have been filtered to an FDR < 5% and an interaction: term size ratio of > 0.2. A comparable enrichment analysis for the corresponding target proteins is provided in Supplementary Table S10.

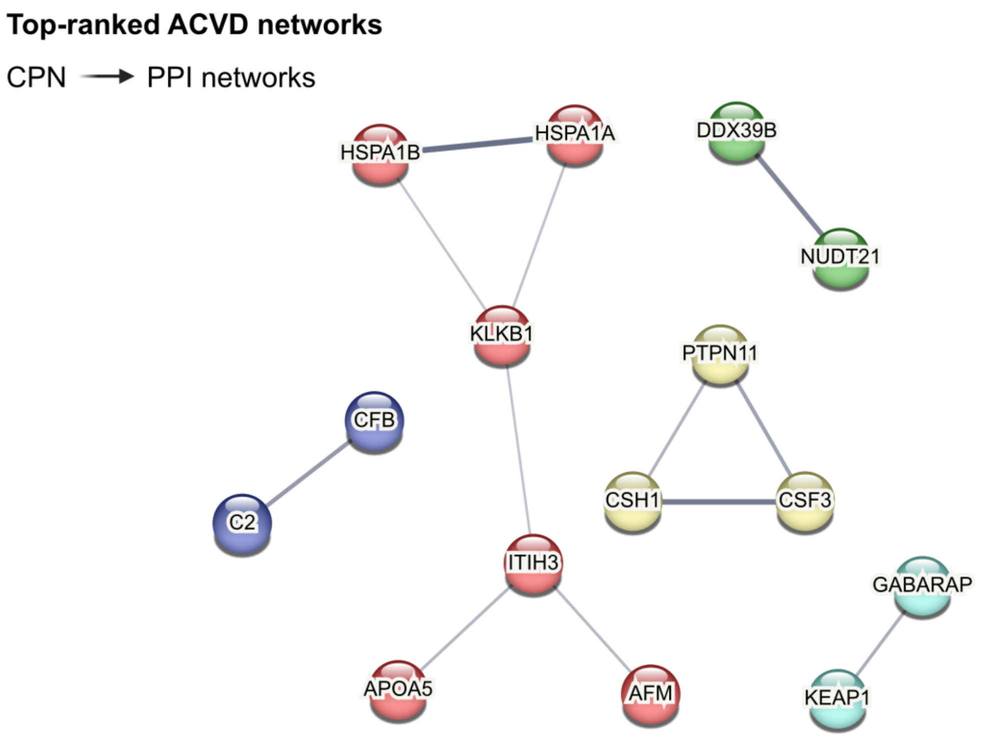


**Supplementary Figure S13**. Protein-protein interactions among network regulators within the top-ranked CPN subnetworks, as identified by the STRING database[9]. These edges represent functional and physical interactions, and unconnected network regulators are excluded from the visualization.
